# Health-related quality of life in children and adolescents born very preterm and its correlates: a cross-sectional study

**DOI:** 10.1101/2024.02.29.24303539

**Authors:** Sarah R Haile, Gabriela P Peralta, Mark Adams, Ajay N Bharadwaj, Dirk Bassler, Alexander Moeller, Giancarlo Natalucci, Thomas Radtke, Susi Kriemler

**Author notes:** Address correspondence to: Susi Kriemler, Epidemiology, Biostatistics and Prevention Institute (EBPI), University of Zurich, Hirschengraben 84, 8001 Zurich, Switzerland.

## Abstract

**Objective:** We aimed to assess health-related quality of life (HRQOL) in a cohort of very preterm born children and adolescents (aged 5-16), and to compare it with their fullterm born siblings and the general population. We also explored correlates of HRQOL among the very preterm born.

**Design:** Cross-sectional survey

**Patients:** Children born <32 weeks gestation (N = 442) as well as their fullterm born siblings (N = 145)

**Main outcome measures:** Primary outcome was KINDL total score (0 worst - 100 best), a validated multidimensional measure of HRQOL in children and adolescents.

**Methods:** Linear mixed models accounted for family unit. Secondary analysis compared very preterm born children to another cohort of healthy children from the same time period. A classification tree analysis explored potential correlates of HRQOL.

**Results:** On average, preterm children, both <28 and 28-31 weeks gestational age, had similar KINDL total score to fullterm sibling controls (-2.3, 95% CI -3.6 to -0.6), and to population controls (+1.4, 95% CI 0.2 to 2.5). Chronic non-respiratory health conditions (such as attention deficit hyperactivity disorder (ADHD) or heart conditions, but not including cerebral palsy), age, and respiratory symptoms affecting daily life were key correlates of HRQOL among very preterm born children.

**Conclusions:** Very preterm birth in children and adolescents was not associated with a relevant reduction in HRQOL compared to their fullterm born peers. However, lower HRQOL was explained by other factors, such as older age, and the presence of chronic non-respiratory health conditions, but also by possibly modifiable current respiratory symptoms. The influence of respiratory symptom amelioration and its potential influence on HRQOL needs to be investigated further.

**What is already known on this topic:** As infants born very preterm become more likely to survive, the importance of health-related quality of life (HRQOL) increases. Research on HRQOL in very preterm born children and adolescents often focuses on non-modifiable risk factors without potential interventions.

**What this study adds:** HRQOL in very preterm born children and adolescents is similar to that of their siblings and to the general population. Age, respiratory symptoms, and chronic health conditions were associated with HRQOL. Better control of respiratory symptoms could improve HRQOL in very preterm born children and adolescents.

**How this study might affect research, practice or policy:** A better understanding of the complex picture of pulmonary disease following prematurity throughout life and interventions to treat respiratory symptoms may be leveraged to improve HRQOL as very preterm born children and adolescents grow.

## Introduction

Recent decades have seen an increased prevalence of very preterm birth (<32 weeks gestation)^1^. These infants are born more and more premature but also increasingly likely to survive the neonatal period^2^. While much research on school-age children born very preterm has focused on neurodevelopment or somatic disease^3^, outcomes such as mental health and health-related quality of life (HRQOL) are of similar importance. It has been suggested that very preterm born children have lower HRQOL than their fullterm counterparts throughout childhood^4,5^, but that these differences do not necessarily persist through adolescence or adulthood^4,6,7^. Some research has specifically focused on HRQOL in preterm children with chronic health conditions, where HRQOL was even found to be similar or only slightly lower than that of their fullterm counterparts^8^. Yet, the many systematic reviews^4,5,7,8^ on HRQOL in the premature born often cannot account for important potentially modifiable factors, and are likely to suffer from inadequate comparison groups or selection bias, including but not limited to selective dropout.

Identifying modifiable correlates of HRQOL is essential in order to develop targeted interventions for HRQOL in the very preterm born. Studies of such correlates with HRQOL have identified factors as gender, maternal education, socio-economic status, nationality^5,9–11^, motor, cognitive or neurodevelopment impairment^6,12,13^, and also behavioral or non-adaptive coping difficulties^12,14^. Although a wide range of potential correlates have been explored previously, few of them are modifiable. Accordingly, it has been recommended that studies exploring long-term outcomes in the preterm born examine lifestyle factors, such as physical activity and diet, and other modifiable factors which could be leveraged to improve HRQOL in this population^15^. In this study, we aimed to compare HRQOL in very preterm born school-age children (<32 weeks gestation) to that of control fullterm siblings (37 weeks or longer), as well as to a population-based cohort from the same time and geographic region. Further, we examined possible correlates, both modifiable and non-modifiable, of HRQOL, using conditional inference trees.

## Methods

In the cross-sectional study FLiP (“Frühgeborenen Lungen Projekt” / Premature Infant Lung Project)^16^, children born less than 32 weeks gestation between January 2006 and December 2019, in the greater Zurich area, Switzerland were recruited. They were all included in the Swiss Neonatal Network & Follow-Up Group (SwissNeoNet), a nationwide registry of very preterm children^17^. Parents of 1401 of 1720 potentially eligible children with valid postal addresses were invited (May - December 2021) to complete an online survey for their preterm child as well as for a term born (37 weeks gestation or later) sibling aged 1 to 18 years, referred as controls hereafter. Families who did not complete the survey within 2 weeks received a reminder call or a second invitation letter, if the phone number was not available. They could also complete a paper version and the questionnaire was available in German, English, French, and Italian. Our analysis included those participants who were at least 5 years of age or older. The study was approved by the Ethics Committee of the Canton of Zurich, Switzerland (2020– 02396). Filling out the online survey was considered as providing consent. The FLiP study was powered to assess the prevalence of respiratory symptoms among children born <32 weeks gestation.

As an additional comparison to schoolchildren from the general population, we used data from the Ciao Corona study, which was part of the Swiss-wide research network Corona Immunitas^18,19^. Ciao Corona was a school-based cohort of randomly selected public and private schools and classes in the canton of Zurich, Switzerland. With 1.5 million inhabitants, the canton of Zurich is largest of 26 cantons in Switzerland by population and is home to a linguistically and ethnically diverse population in both urban and rural settings. While the primary endpoint of Ciao Corona was seropositivity, questionnaires included a range of other measures, including the KINDL^20^, assessed repeatedly between June 2020 and December 2022. For comparison with FLiP, the KINDL total score from September 2021 was used, as this best matched the timeframe of the FLiP assessment period. The Ciao Corona study was approved by the Ethics Committee of the Canton of Zurich, Switzerland (2020-01336). All participants provided written informed consent before being enrolled in the study.

The primary outcome was the KINDL total score^21,22^, a validated instrument for assessing HRQOL ranging from 0 (worst) to 100 (best) (for further details, see Supplementary Material and Table S1). Secondary outcomes included all the KINDL subscales (physical, emotional, self-esteem, family, friends, and school). Additional data collected included participants’ age and gender, gestational age (in weeks, range 24 - 31), birthweight (in grams), diagnosed bronchopulmonary dysplasia (BPD), socio-economic status (SES), family unit, chronic health conditions, hours of physical activity per week, hours of screen time per week, participation in music lessons, participation in scouts, participation in sports, and need for various types of therapy. Prematurity-related diagnosis of BPD was taken from personal history of the premature born children included in the SwissNeoNet registry (none to mild vs moderate to severe). SES was determined according to each parent’s education level (1 university, 2 vocational university, 3 apprenticeship, 4 job requiring minimal training, 5 compulsory education, 6 less than compulsory education), and then summed over both parents (range 2 highest education - 12 lowest education). Chronic health conditions were categorized as respiratory, non-respiratory or cerebral palsy. Respiratory conditions included asthma and cystic fibrosis. Non-respiratory conditions included heart conditions, diabetes, intestinal issues, low/high blood pressure, attention deficit hyperactivity disorder, epilepsy, joint disorders, and depression. Cerebral palsy was reported separately along with its severity (none; mild, no to minimal restriction to daily activities; mild, limitations in daily activities but without the need for aids; moderate, needs prostheses, medication or technical aids to manage daily activities; severe, requires a wheelchair and has significant difficult in daily activities). Types of therapy included speech, physical, occupational, psychomotor, curative or psychological therapy, as well as early support programs. To assess whether respiratory symptoms affected daily life, parents were asked about several questions related to whether their child had cough or wheezing due to physical exertion in the last 12 months or whether cough, or wheezing restricted their daily activities. Other included variables were: number of siblings, presence of house pets, whether parents smoked (no/outside/in the home), number of therapies used, use of assistive devices (e.g hearing aids, walking aids, wheelchair), hours of physical activity per day, hours of screen time per day, and participation in sports, scouts, or musical activities (see Supplementary Material for wording of selected questions).

Key demographic variables were summarized as median (range), n (%), or in the case of socio-economic status, median [IQR]. Outcomes were compared between FLiP preterm and FLiP control participants using linear mixed models, including family unit as a random effect. Comparisons of FLiP preterm and Ciao Corona control participants were made using linear regression, after 2:1 matching on age in years, sex and nationality. Sensitivity analyses included a) excluding participants with chronic health conditions, b) restricting to preterm born children with control siblings, c) stratification by age, d) adjusting for SES, and e) using fixed effects to account for family unit. Coefficients and corresponding 95% confidence intervals were interpreted according to their possible relevance, rather than with p-values^23,24^. To explore other potential correlates, both modifiable and non-modifiable, of HRQOL among very preterm born children, we used conditional inference trees^25,26^ estimated by binary recursive partitioning. To handle missing predictor values, the conditional inference trees used up to 3 surrogate splits^25^. The algorithm stopped if no split with *α* < 0.05 could be constructed or if a subgroup had less than 25 participants. For further details, see Supplementary Material.

The statistical analysis was performed using R (R version 4.4.1 (2024-06-14)). Linear mixed models were fit using the `lmerTest` package^27^, and tables were produced with `gtsummary`^28^. The classification trees were fit using the `ctree` function from `partykit`^29^. Nearest neighbor matching using robust rank-based Mahalanobis distance to the Ciao Corona data was performed using the `MatchIt` package^30^.

### Patient and Public Involvement

The FLiP survey used input from parents to optimize content and clarify the questionnaire. A small group of very preterm born children and adolescents as well as members of the public tested the questionnaires in a pilot phase, and were given the opportunity to comment on patient information leaflets.

In the Ciao Corona study, some school principals were consulted during the development of the protocol to ensure feasibility of the planned study procedures. Feedback from invited and enrolled children and parents was continuously collected to adapt the communication strategies and channels. Children and parents always received their individual serological results with interpretation. Regular fact sheets were sent to participants and the public. Online information sessions were organised at the beginning, middle and end of the study to encourage open exchange and feedback for invited and enrolled school principals, staff, and parents of the children.

## Results

After inviting 1401 very preterm born children and their parents to participate in FLiP, data from 681 40% preterm children and 205 fullterm control siblings was available. Among the very preterm born, we excluded 199 children that were younger than 5 years of age and 40 had not filled out KINDL, leaving 442 preterm participants. Among their fullterm born siblings, 56 were younger than 5 years of age and 4 had not filled out KINDL, leaving 145 fullterm siblings (see **Supplementary Figure S1**). We also included 1058 participants from Ciao Corona (total n=4435, 2020 - 2022, of whom 2974 had participated prior to September 2021^31^) with a KINDL total score in September 2021. Characteristics of the included participants are found in **Table 1**, and not included participants were similar regarding most morbidities typical for the very preterm (**Supplementary Table S2**).

**Table 1:** Key characteristics of FLiP very preterm born children and adolescents, their control siblings, and age, sex and nationality matched control participants from Ciao Corona. BPD indicates bronchopulmonary dysplasia. Chronic health conditions included asthma, cystic fibrosis, congenital heart defects, heart disease, celiac, diabetes, inflammatory bowel disease, high blood pressure, attention deficit hyperactivity disorder, epilepsy, joint disorders, depression/anxiety, and cerebral palsy. Socio-economic status is measured on the basis of parents’ education from 2 (both parents having university education) to 12 (both parents less than compulsory education) points, though the categories differed somewhat in FLiP and Ciao Corona. As physical activity was assessed using different questions in Ciao Corona than in FLiP that were not comparable, we did not include physical activity in Ciao Corona here.

| Characteristic | FLiP preterm N = 442 | FLiP control N = 145 | Ciao Corona control N = 882 |
| --- | --- | --- | --- |
| age (years) | 10 (5 - 16) | 9 (5 - 19) | 10 (6 - 16) |
| sex (male) | 236 (53%) | 84 (59%) | 472 (54%) |
| gestational age (weeks) |  |  |  |
| 24-27 wks | 130 (29%) |  |  |
| 28-31 wks | 312 (71%) |  |  |
| birthweight |  |  |  |
| <1000g | 158 (36%) |  |  |
| 1000+g | 284 (64%) |  |  |
| multiple gestation | 140 (32%) |  |  |
| socio-economic status | 5 [3, 6] | 5 [3, 6] | 4 [3, 5] |
| non-Swiss nationality | 66 (15%) | 20 (14%) | 121 (14%) |
| moderate to severe BPD | 55 (12%) |  |  |
| coughing / wheezing restrict daily activities | 13 (2.9%) | 1 (0.7%) |  |
| any chronic health condition | 104 (24%) | 12 (8.3%) | 122 (14%) |
| chronic non-respiratory conditions | 67 (15%) | 11 (7.6%) | 96 (11%) |
| chronic respiratory conditions | 24 (5.4%) | 3 (2.1%) | 34 (3.9%) |
| cerebral palsy | 33 (7.5%) | 1 (0.7%) | 0 (0%) |
| physical activity (hours per day) | 0.71 [0.50, 1.00] | 0.71 [0.57, 1.14] |  |

<sup>1</sup> Median (Range); n (%); Median [IQR]

Very preterm born children with gestational age 24-27 weeks (n = 130) had an average total KINDL score of 77.3 ± 10.0 points out of 100, while those with gestational age 28-31 weeks (n = 312) had an average total KINDL score of 78.9 ± 10.5, compared to 80.8 ± 8.7 among fullterm born control siblings. On average, preterm children born at 24-27 weeks had a 2.3 point lower KINDL total score than fullterm controls (95% CI -4.4 to -0.2), when accounting for family unit. Similarly, those born at 28 weeks had a 2.3 point lower KINDL total score than fullterm controls (95% CI -3.9 to -0.7) (**Figure 1, Supplementary Table S3**). The pattern was similar for the KINDL subscales (**Supplementary Figure S2**), and when comparing those with birthweight <1000g with those at least 1000g (**Supplementary Figures S3 and S4, Supplementary Table S4**). Results were similar when examining all preterm born children together for both KINDL total score and its subscales (**Supplementary Figures S5 and S6, Table S5**), and did not change in any of the sensitivity analyses.

**Figure 1:**
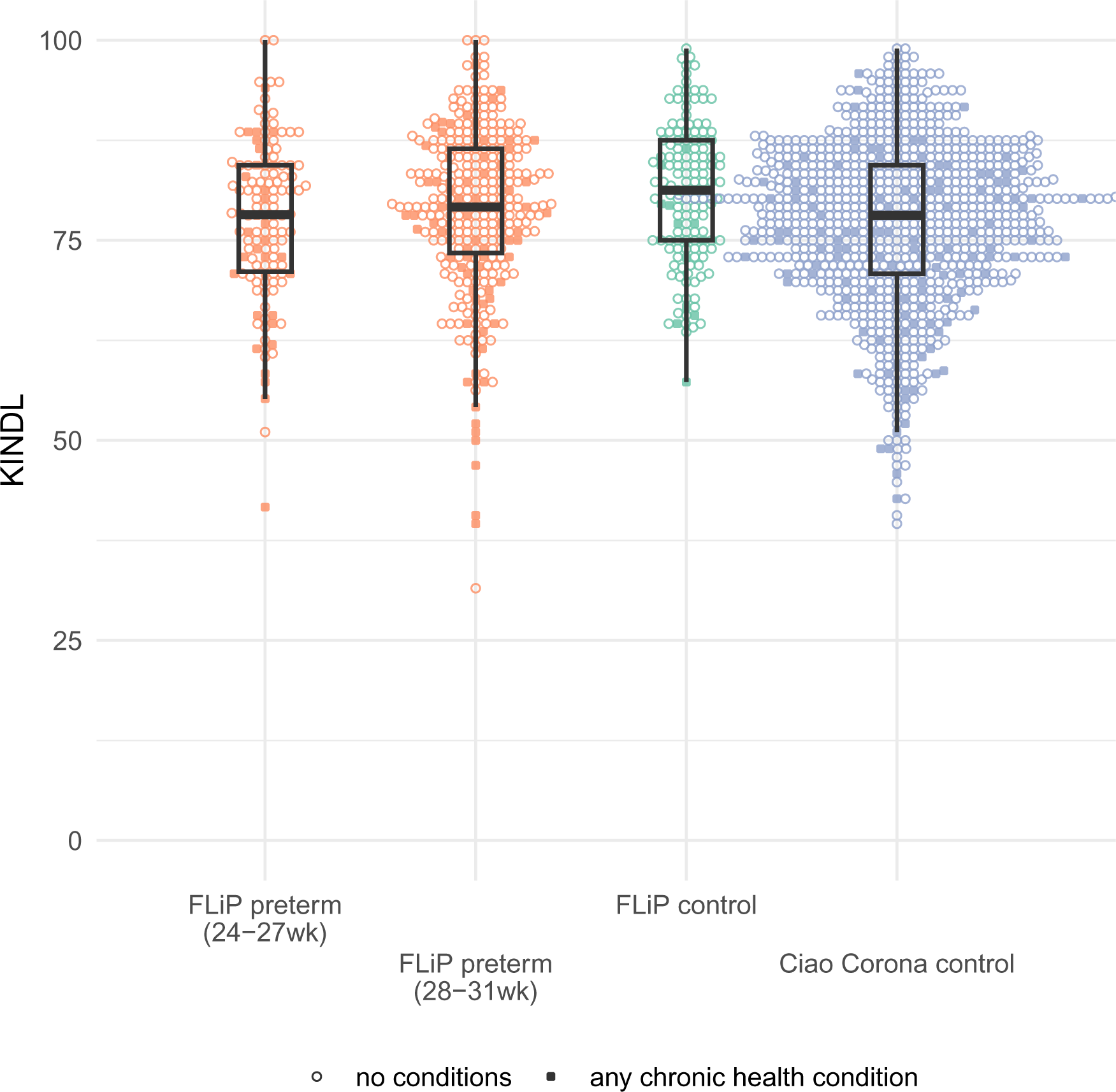
KINDL total score, for very preterm born children (FLiP preterm, stratified by gestational age 24-27 weeks or 28-31 weeks) and their fullterm born siblings (FLiP control), as well as age, sex and nationality-matched participants from Ciao Corona (Ciao Corona control). Solid circles indicate participants without chronic health conditions, while empty diamonds indicate those with any chronic health condition, respiratory or non-respiratory.

A number of other potential non-modifiable and modifiable correlates for HRQOL were further considered in a classification tree analysis (**Supplementary Table S6**). It indicated that respiratory symptoms affecting daily activities as well as age and chronic non-respiratory conditions were the primary correlates of HRQOL among very preterm born children and adolescents in our sample (**Figure 2**), with mean KINDL total score ranging from 68.2 ± 13.1 (among those 10 years of age or older with chronic non-respiratory conditions) to 80.5 ± 8.8 (among those with no chronic health conditions and no respiratory symptoms affecting daily activities). 67% (296 / 442) of the sample was in the latter group that could not be further differentiated with the selected variables. Notably, gestational age, birthweight and BPD were not identified as correlates of HRQOL in our sample.

**Figure 2:**
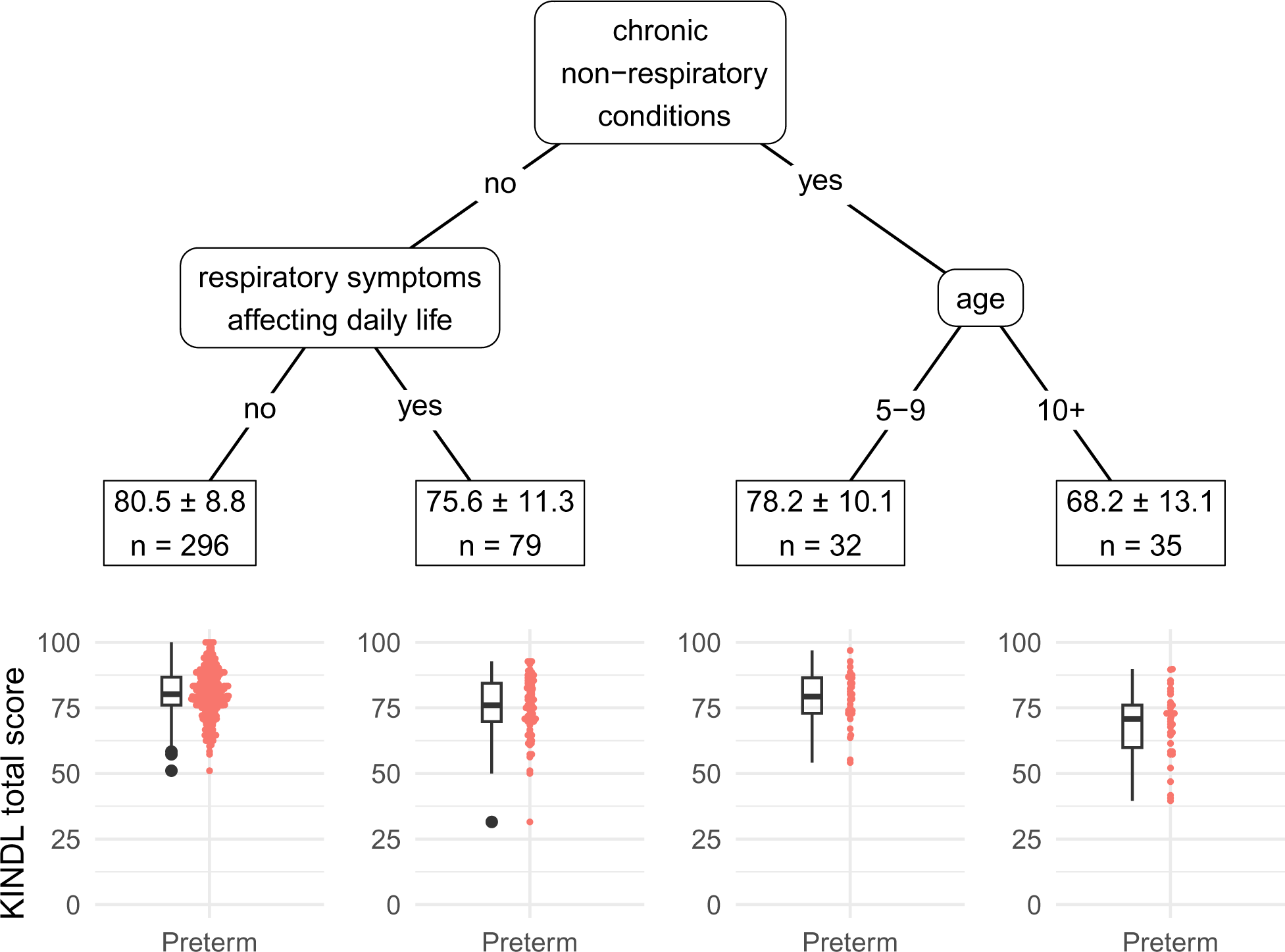
Classification tree for KINDL total score in very preterm born children, based on a range of possible correlates (see Table S7). Identified correlates are chronic non-respiratory conditions, age group (5-9 vs 10+), and whether respiratory symptoms negatively affect daily life. For each identified subgroup, mean ± standard deviation for KINDL total score is given, along with the number of participants.

## Discussion

We observed no relevant difference in HRQOL (KINDL total score) when comparing very preterm children to their fullterm siblings, even after accounting for gestational age or birth-weight, chronic health conditions, including respiratory conditions or cerebral palsy, and did not change in any of the sensitivity analyses. HRQOL among very preterm children was also similar to that of the general population of schoolchildren. When considering possible variables associated with HRQOL beyond prematurity, respiratory symptoms affecting daily activities, chronic non-respiratory conditions, and age group appeared to play a role and were identified as more important correlates of HRQOL than gestational age. Two points out of 100 difference in the total KINDL score represent a marginal difference in HRQOL that was not clinically relevant. Comparing KINDL in children with and without various chronic health conditions, differences ranging from 1.9 (children with asthma) to 6.2 (cancer survivors) points have been observed^32–34^. The 2 point difference in our study was thus quite small, and based on the variability of the KINDL total score (SD=10.3), likely not meaningful.

In a classification tree analysis the most important factors correlated with HRQOL appeared to be age group, chronic health conditions and the existence of respiratory symptoms affecting daily activities. While age and chronic health conditions are not modifiable, the presence of respiratory symptoms may be modifiable, indicating that improvement of respiratory symptoms may potentially improve HRQOL in the very preterm born.

Children with respiratory symptoms should be investigated for their phenotype profile and potentially treatable traits by a pediatric pulmonologist to understand the complex picture of prematurity associated respiratory symptoms, and whether or not the child may benefit from treatment at all or treatment optimization^35^. This is important as up to 40% of the premature population are prescribed asthma medication during childhood^35^, although there is a lack of objective evidence on how to treat these individuals and whether treatment improves symptoms^35–37^.

Thus, individual treatment needs to be based on the phenotype or underlying mechanisms of prematurity-associated lung disease as proposed by for instance the wheel-and-spoke model that combines components of a phenotype classification including structural, physiological, inflammatory and clinical traits^35^. Non-pharmaceutical interventions such as exercise to improve cardiopulmonary function might likewise be of benefit^38^. Our finding that respiratory symptoms are correlated with HRQOL could imply that interventions targeting those symptoms may potentially also improve HRQOL in the very preterm born. This hypothesis of course should be tested in future studies.

Our analysis has several strengths. HRQOL was assessed using the validated^22^ multidimensional KINDL instrument for children and adolescents. The analysis used a relatively large registry of very preterm born children in Switzerland and included fullterm born siblings as a control group. Family unit was accounted for in the analysis. The Ciao Corona study provided a school-based random sample of school children in the same geographic region and time period. To account for differing severity in terms of prematurity, we stratified the analysis by gestational age and birthweight. We considered a broad range of possible correlates, including respiratory symptoms, of HRQOL in a classification tree analysis, which has to our knowledge not been performed previously in this population, and allowed us to explore a wide range of possible correlates with HRQOL.

A key limitation is that longitudinal data on HRQOL was not available for this sample of children, and not all very preterm born participants had control siblings. We did not have information on other potentially important variables, for example, on participants’ mental health status (e.g. sadness or anxiousness^39^) or on social support^40^, which could have provided useful information in the conditional inference tree analysis. There were also not many fullterm born children with chronic health conditions in the FLiP sample, which would have allowed us to further explore the associations between very preterm birth, chronic health conditions and HRQOL. Like other studies of very preterm born children, our sample may have had selection bias^5,7^. Although this research took place during the COVID-19 pandemic which may have affected HRQOL of the children, but if so, this was likely true for all included groups. Nevertheless, we cannot exclude that premature born children were especially shielded, which would compromise HRQOL even more. Yet, this was not the case and HRQOL was similar in normal school children and previously premature children.

It is a gift of medicine that children born <32 weeks gestation generally have a HRQOL comparable to fullterm born children^6^. Nevertheless, as observed in this large cohort of very preterm born children, there are children that clearly show compromised HRQOL. Low HRQOL was not restricted to those born prior to 28 weeks of gestational age or less than 1000g birthweight, as seen in our stratified analyses. While an association between HRQOL and older age or chronic health conditions may often be expected and considered plausible, the association with respiratory symptoms observed in our analysis may be neglected and often not addressed^41^.

A better understanding of the complex picture of pulmonary disease following prematurity throughout life and interventions to treat respiratory symptoms, such as medical treatment or those targeting aerobic exercise and physical activity, may be leveraged to improve HRQOL as very preterm born children and adolescents grow.

## Data Availability

Deidentified individual participant data (including data dictionaries) will be made available, in addition to study protocols, the statistical analysis plan, and the informed consent form. The data will be made available upon publication to researchers who provide a methodologically sound proposal for use in achieving the goals of the approved proposal. Proposals should be submitted to Susi Kriemler.

## Acknowledgments

The authors thank Agnė Ulytė, Alessia Raineri and Sonja Rüegg (all University of Zurich, Switzerland) for their work on data collection for the Ciao Corona study.

## Ethical approval

The FLiP study was approved by the Ethics Committee of the Canton of Zurich, Switzerland (2020–02396). Filling out the online survey was considered as providing consent. The Ciao Corona study was approved by the Ethics Committee of the Canton of Zurich, Switzerland (2020-01336). All participants provided written informed consent before being enrolled in the study.

## Role of Funder/Sponsor (if any)

The funding sources had no role in the design and conduct of the study; collection, management, analysis, and interpretation of the data; preparation, review, or approval of the manuscript; and decision to submit the manuscript for publication.

## Clinical Trial Registration (if any)

Ciao Corona was registered at ClinicalTrials.gov: NCT04448717.

## Abbreviations

BPD: Bronchopulmonary dysplasia
FLiP: “Frühgeborenen Lungen Projekt” / Premature Infant Lung Project
HRQOL: health-related quality of life
SES: Socio-economic status

## Contributors Statement

Sarah R Haile, PhD conceptualized and designed the study, conducted the statistical analysis, drafted the initial manuscript, and critically reviewed and revised the manuscript.

Gabriela P Peralta, PhD acquired funding, collected and cleaned the data and critically reviewed and revised the manuscript.

Mark Adams, PhD, Dirk Bassler, MD, Alexander Moeller, MD, and Giancarlo Natalucci, MD acquired funding, collected data and critically reviewed and revised the manuscript.

Ajay N Bharadwaj, BSc collected data and critically reviewed and revised the manuscript.

Thomas Radtke, PhD acquired funding, conceptualized and designed the study, and critically reviewed and revised the manuscript for important intellectual content.

Susi Kriemler, MD acquired funding, conceptualized and designed the study, coordinated and supervised data collection, critically reviewed and revised the manuscript for important intellectual content, is responsible for the overall content as guarantor.

All authors approved the final manuscript as submitted and agree to be accountable for all aspects of the work.

