## Supplemental Table for "Health-related quality of life in children and adolescents born very preterm and its correlates: a cross-sectional study"

**Computational Details** **31**

#### List of Tables

S1 KINDL questions as used in the FLiP and Ciao Corona studies. Item are noted with an asterisk (*) if the wording has been adapted for FLiP. . . . . 5

S2 Comparisons of very preterm born children included in this analysis ('analyzed') vs not included in this analysis. IVH indicates intraventricular hemorrhage, PVL periventricular leukomalacia, NEC necrotizing enterocolitis, ROP retinopathy of prematurity, and NDI neurodevelopmental impairment. . . . . 10

S3 Differences in KINDL total score between very preterm born children and their fullterm siblings from FLiP or participants from Ciao Corona, stratified by gestational age (24-27 weeks vs 28-31 weeks). Mean differences denote either differences in health-related quality of life between FLIP very preterm born siblings and their control siblings, or between FLIP very preterm born siblings and controls from Ciao Corona. Negative mean differences indicate that very preterm born children had lower HRQOL than controls. Mean differences and 95% confidence intervals account for either family unit or matching and therefore may not strictly correspond to the means for each group given in the first and second columns. . . . . 11

S4 Differences in KINDL total score between very preterm born children and their fullterm siblings from FLiP and participants from Ciao Corona, stratified by birthweight ($<$ 1000g vs $\geq$ 1000g). Mean differences denote either differences in health-related quality of life between FLIP very preterm born siblings and their control siblings, or between FLIP very preterm born siblings and controls from Ciao Corona. Mean differences and 95% confidence intervals account for either family unit or matching and therefore may not strictly correspond to the means for each group given in the first and second columns. . . . . 13

S5 Differences in KINDL total score between very preterm born children and their fullterm siblings from FLiP and participants from Ciao Corona. Mean differences denote either differences in health-related quality of life between FLIP very preterm born siblings and their control siblings, or between FLIP very preterm born siblings and controls from Ciao Corona. Mean differences and 95% confidence intervals account for either family unit or matching and therefore may not strictly correspond to the means for each group given in the first and second columns. . . . . 16

S6 Table of potential determinants of health-related quality of life in FLiP among very preterm born children and adolescents, by age group. . . . . 19

S7 Differences in KINDL total score between very preterm born children, their fullterm born siblings, and Ciao Corona participants (stratified by presence of any chronic health conditions). In FLiP, 104 of very preterm born children had chronic health conditions (24%), while n = 12 of their control siblings did (8%). Among controls from Ciao Corona, 122 (14%) reported chronic health conditions. Mean differences denote either differences in health-related quality of life between FLIP very preterm born siblings and their control siblings, or between FLIP very preterm born siblings and controls from Ciao Corona. Mean differences and 95% confidence intervals account for either family unit or matching and therefore may not strictly correspond to the means for each group given in the first and second columns. . . . . 21

S8 Differences in KINDL total score between preterm children and their fullterm siblings (only matched siblings). . . . . 23

S9 Key characteristics of preterm and control participants in full data (preterm children with included control siblings) . . . . . 24

S10 Differences in KINDL total score between very preterm born children and their fullterm siblings from FLiP and participants from Ciao Corona, by age group (5-9, 10+). Mean differences, 95% confidence intervals and p-values account for either family unit or matching and therefore may not strictly correspond to the means for each group given in the first and second columns. . . . 25
S11 Differences in KINDL total score between very preterm born children and their fullterm siblings from FLiP. Results from models accounting for family unit as 1) random effect, or 2) fixed effect are shown. Mean differences and 95% confidence intervals are shown. . . . . . . . . . . . . . 29
S12 Differences in KINDL total score between very preterm born children and their fullterm siblings from FLiP, by gestational age. Results from models accounting for family unit as 1) random effect, or 2) fixed effect are shown. Mean differences and 95% confidence intervals are shown. 30

#### Supplementary Methods

##### The KINDL score

The KINDL-R score (hereafter, KINDL) is a validated instrument for measuring health-related quality of life} [1], with scores ranging from 0 (worst) to 100 (best). In FLiP, we used slightly adapted versions of the parent (proxy) versions for 4-6 year olds and for 7-17 year old children and adolescents (https://www.kindl.org/contacts/english/). In Ciao Corona, we used the parent (proxy) version, for 7-17 year old children and adolescents. All questions as asked are listed in Table S1. The KINDL contains 24 items, 4 in each of 6 subscales: physical well-being, emotional well-being, self-esteem, family, social contacts / friends, and school. To compute the KINDL score, certain items are first recoded. A subscale can be analysed as long as no more than 30% of its items are missing. Mean value replacement is used to deal with missing item scores. The 6 subscales are then added and rescaled to 0-100 to form the KINDL total score. Sample code is available at https://www.kindl.org/english/analysis/. Text in English for each of the items is provided below, but FLiP and Ciao Corona used German versions of the questionnaires.

**Table S1:** KINDL questions as used in the FLiP and Ciao Corona studies. Item are noted with an asterisk (*) if the wording has been adapted for FLiP.

| item | FLiP (age 3 - 6 years) | FLiP (age 7 – 17 years) | Ciao Corona |
|---|---|---|---|
| **1. Physical Well-being** | | | |
| | During the past week... | During the past week... | During the past week... |
| 1 | ... my child felt ill. | ... my child felt ill. | ... my child felt ill. |
| 2 | ... my child had a headache or tummyache. | ... my child had a headache or tummyache. | ... my child had a headache or tummyache. |
| 3 | ... my child my child was tired and worn-out. | ... my child my child was tired and worn-out. | ... my child my child was tired and worn-out. |
| 4 | ... my child felt strong and full of energy. | ... my child felt strong and full of energy. | ... my child felt strong and full of energy. |
| **2. Emotional Well-being** | | | |
| | During the past week... | During the past week... | During the past week... |
| 1 | ... my child had fun and laughed a lot | ... my child had fun and laughed a lot | ... my child had fun and laughed a lot |
| 2 | ... my child didn't feel much like doing anything | ... my child didn't feel much like doing anything | ... my child didn't feel much like doing anything |
| 3 | ... my child felt alone | ... my child felt alone | ... my child felt alone |
| 4 | ... my child felt alone | ... my child felt alone | ... my child felt alone |
| **3. Self-esteem** | | | |
| | During the past week... | During the past week... | During the past week... |
| 1 | ... my child was proud of him-/herself | ... my child was proud of him-/herself | ... my child was proud of him-/herself |
| 2 | ... my child felt on top of the world | ... my child felt on top of the world | ... my child felt on top of the world |
| 3 | ... my child felt pleased with him-/herself | ... my child felt pleased with him-/herself | ... my child felt pleased with him-/herself |
| 4 | ... my child had lots of good ideas | ... my child had lots of good ideas | ... my child had lots of good ideas |
| **4. Family** | | | |
| | During the past week... | During the past week... | During the past week... |
| 1 | ... my child got on well with us as parents | ... my child got on well with us as parents | ... my child got on well with us as parents |
| 2 | ... my child felt fine at home | ... my child felt fine at home | ... my child felt fine at home |
| 3 | ... we quarrelled at home | ... we quarrelled at home | ... we quarrelled at home |
| 4 | ... my child felt that I was bossing him/her around | ... my child felt that I was bossing him/her around | ... my child felt that I was bossing him/her around |
| **5. Social Contacts** | | | |

**Table S1:** KINDL questions as used in the FLiP and Ciao Corona studies. Item are noted with an asterisk (*) if the wording has been adapted for FLiP. *(continued)*

| item | FLiP (age 3 - 6 years) | FLiP (age 7 – 17 years) | Ciao Corona |
|---|---|---|---|
| | During the past week... | During the past week... | During the past week... |
| 1* | ... my child played or did things together with friends | ... my child played or did things together with friends | ... my child did things together with friends |
| 2 | ... my child was liked by other kids | ... my child was liked by other kids | ... my child was liked by other kids |
| 3 | ... my child got along well with his/ her friends | ... my child got along well with his/ her friends | ... my child got along well with his/ her friends |
| 4 | ... my child felt different from other children | ... my child felt different from other children | ... my child felt different from other children |
| **6. School** | | | |
| | During the past week... | During the past week... | During the last week in which my child was at school ... |
| 1* | ... my child coped well with the assignments set in nursery school/ kindergarten | ... my child easily coped with schoolwork | ... my child easily coped with schoolwork |
| 2* | ... my child enjoyed the nursery school/ kindergarten | ... my child enjoyed the school lessons | ... my child enjoyed the school lessons |
| 3* | ... my child looked forward to nursery school/kindergarten | ... my child worried about his/her future | ... my child worried about his/her future |
| 4* | ... my child made lots of mistakes when doing minor assignments or homework | ... my child was afraid of bad marks or grades | ... my child was afraid of bad marks or grades |

#### Chronic health conditions listed on the surveys

- **FLiP**: asthma, cystic fibrosis, congenital heart defects, heart disease, celiac / gluten allergy , lactose intolerance, allergies (other than hay fever), diabetes mellitus, chronic inflammation of the bowel (ulcerative colitis or Crohn's disease), high blood pressure (hypertension), attention deficit disorder (ADHD, ADD), epilepsy, joint disease (e.g. arthritis), depression/anxiety disorder, other [comment field], cerebral palsy [with severity]
- **Ciao Corona**: asthma, hay fever, celiac, lactose intolerance, allergies (other than hay fever), neurodermatitis / excema, Diabetes Mellitus, chronic inflammation of the bowel (ulcerative colitis or Crohn's disease), high blood pressure (hypertension), attention deficit disorder (ADHD, ADD), epilepsy, joint disease (e.g. arthritis), depression/anxiety disorder, other [comment field]

The following conditions were not counted as chronic health conditions for the purposes of this analysis: hay fever, celiac, gluten allergy, lactose intolerance, allergies (other than hay fever). The Ciao Corona study did not ask specifically about cerebral palsy, but it was also not reported under "other".

#### Selected Questions from FLiP

- **Respiratory symptoms affecting daily life**: Yes to any of the following questions:

  a) *In the last 12 months, has your child ever had whistling or wheezing breathing during or after physical exertion?*
  b) *Have any of the following situations triggered a cough in your child in the last 12 months? Physical exertion (running, sports)*
  c) *Have any of the following situations triggered whistling or wheezing in your child in the last 12 months? Physical exertion (running, sports)*
  d) *Does your child sometimes have difficulty breathing during physical exertion?*
  e) *In the last 12 months, how much was your child's daily activities (or play behaviour) restricted by the cough?*
  f) *In the last 12 months, how much was your child restricted in his/her daily activities (or play behaviour) because of whistling or wheezing breathing or shortness of breath?*

- **Physical Activity**: *On average, how many hours per week does your child spend in physical activity that causes at least some sweating or heavy breathing? (School sport INCLUDED)* This value was converted to hours per day.
- **Screen Time**: These values were converted to average hours per day.

  a) *How many hours per day does your child CURRENTLY spend using electronic devices on a typical weekday? NOT counting school lessons and schoolwork. For example: Mobile phone, tablet, Playstation, Xbox, Nintendo, computer, TV*
  b) *How many hours per day does your child CURRENTLY spend using electronic devices on a typical weekend day? NOT counting school lessons and schoolwork. For example: Mobile phone, tablet, Playstation, Xbox, Nintendo, computer, TV*

- **Sports**: *Does your child participate regularly (at least once every 2 weeks) in the following activities? Yes, sport activities such as gymnastics club, ballet, dance, tennis, basketball, football club etc.*
- **Music**: *Does your child participate regularly (at least once every 2 weeks) in the following activities? Yes, music lessons, musical activities, theatre, circus, etc.*
- **Scouts**: *Does your child participate regularly (at least once every 2 weeks) in the following activities? Yes, other activities such as Scouts, Cevi [YMCA / YWCA], Blauring / Jungwacht [similar Catholic organization], etc.*
- **SES**: Sum of *What is the highest educational qualification of the mother?* and *What is the highest educational qualification of the father?* as defined by SwissNeoNet https://app.swissneonet.ch/data/live/structure/38/show

  1. University
  2. University of applied science, technical college, higher level job training, college of education, university entrance qualification (Matura or Berufsmatura)
  3. Apprenticeship or secondary school diploma
  4. Job requiring minimal training
  5. Regular school without job training
  6. No education, or unfinished regular school

#### Detailed Statistical Methods: Conditional Inference Trees

We used conditional inference trees [2, 3] estimated by binary recursive partitioning to identify possible determinants of health-related quality of life (HRQOL) in very preterm born children and adolescents. These models seek to make homogeneous subgroups, i.e. clusters, of the sample with respect to the outcome of interest. Generally, the algorithm 1) searches for the variable with the strongest association to the outcome, and then 2) splits the values of that variable into two groups, and repeats this process until some stopping criteria (in this analysis, p-value < 0.05 or sample size < 25) are reached. While all types of variables can be selected in step 1 of this algorithm, categorical and continuous variables have more flexibility in terms of selected thresholds in step 2 than binary variables do. Conditional inference trees select variables in an unbiased manner, without being affected by overfitting [2]. Such models identify subgroups defined by combinations of covariates, without needing to *a priori* specify interaction terms or consider multicollinearity [4], and thresholds do not need to be prespecified. Standard regression models, even when combined with model selection, do not identify homogenous subgroups. Due the presence of missing observations in some of the variables, we employed so-called surrogate splits to account for this missingness without excluding subjects [2].

The conditional inference trees were fit using `ctree` from the R package `partykit` [2, 5]. Missing covariate information was handled by `ctree` directly. All analysis was performed in R (R version 4.4.1 (2024-06-14)).

### Supplementary Results

#### Analysis Population

##### Flowchart

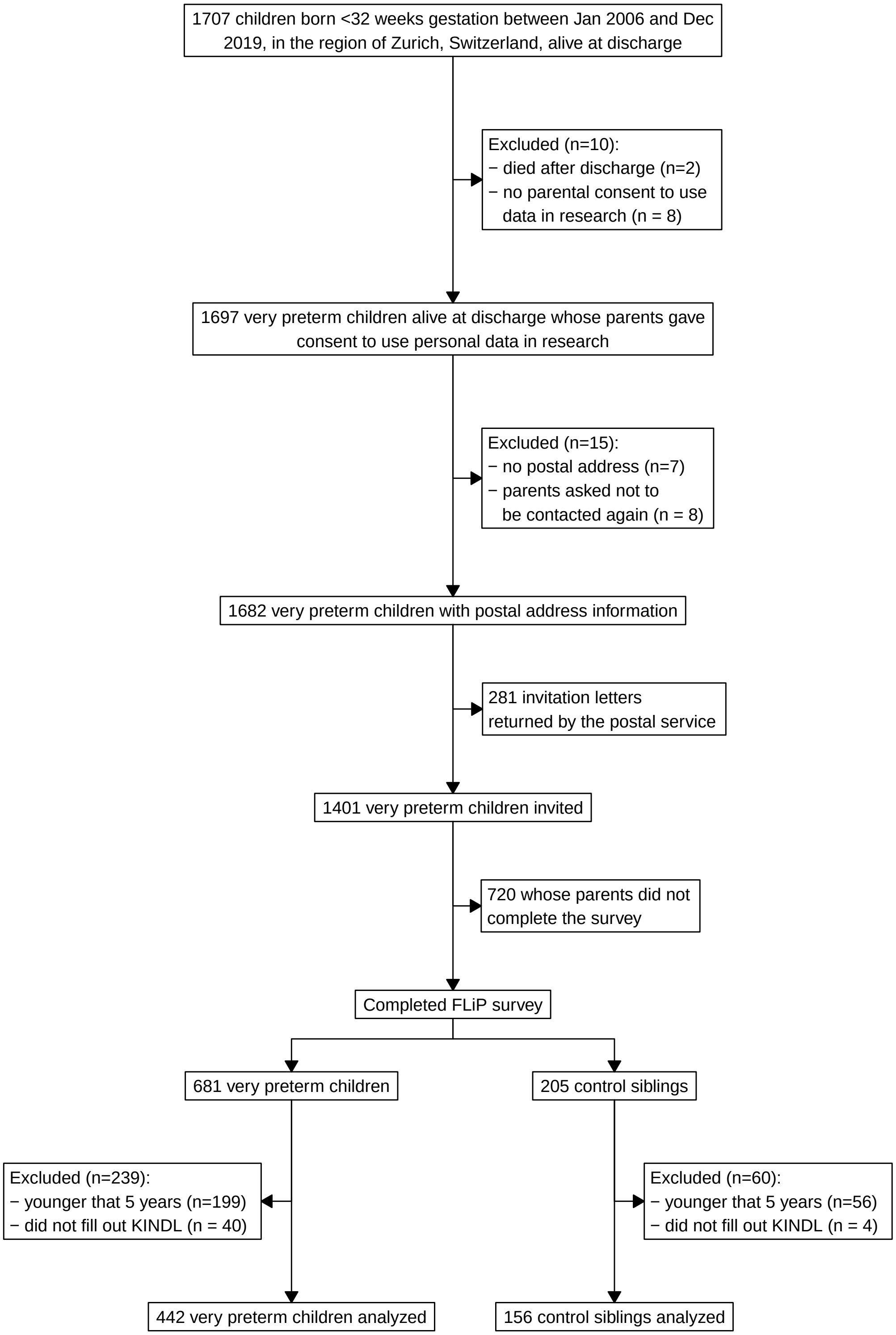

**Figure S1:** Flowchart of children and adolescents included in the FLiP cohort study.

**Analyzed vs not analyzed very preterm born children**

Very preterm born children included in our analysis were generally comparable to those not included, though with slightly lower gestational age and birthweight, and more likely to have had supplemental oxygen at 36 weeks, as moderate to severe BPD is often defined. Notably, none of the other morbidities typical for very preterm children are different between participants and non-participants (**Supplementary Table S2**).

**Table S2:** Comparisons of very preterm born children included in this analysis ('analyzed') vs not included in this analysis. IVH indicates intraventricular hemorrhage, PVL periventricular leukomalacia, NEC necrotizing enterocolitis, ROP retinopathy of prematurity, and NDI neurodevelopmental impairment.

| variable | not analyzed | analyzed | p-value | Total |
|---|---|---|---|---|
| N | 1031 (70%) | 442 (30%) | | 1473 |
| Gestational age (IQR) | 30.6 (28.6 to 31.7) | 29.4 (27.4 to 30.7) | <0.0001 | 30.1 (28.1 to 31.4) |
| Birth weight z-score (Voigt 2006) (IQR) | -0.2 (-1.2 to 0.4) | 0 (-0.6 to 0.4) | <0.0001 | -0.1 (-1 to 0.4) |
| Sex male N (%) | 563 (54.6 %) | 236 (53.4 %) | 0.710 | 799 (54.2 %) |
| Outborn N (%) | 29 (2.8 %) | 9 (2 %) | 0.495 | 38 (2.6 %) |
| Multiple births N (%) | 376 (36.5 %) | 140 (31.7 %) | 0.088 | 516 (35 %) |
| Any antenatal steroids N (%) | 910 (91.6 %) | 397 (92.5 %) | 0.642 | 1307 (91.9 %) |
| Caesarean section N (%) | 889 (86.2 %) | 382 (86.4 %) | 0.985 | 1271 (86.3 %) |
| Congenital malformation (validated) N (%) | 20 (1.9 %) | 7 (1.6 %) | 0.799 | 27 (1.8 %) |
| Severe IVH N (%) | 35 (3.4 %) | 18 (4.1 %) | 0.625 | 53 (3.6 %) |
| Cystic PVL N (%) | 9 (0.9 %) | 4 (0.9 %) | 1.000 | 13 (0.9 %) |
| Supplemental oxygen at 36 weeks GA* N (%) | 72 (7 %) | 55 (12.4 %) | 0.001 | 127 (8.6 %) |
| NEC stage >=2 N (%) | 16 (1.6 %) | 7 (1.6 %) | 1.000 | 23 (1.6 %) |
| Severe ROP* N (%) | 21 (2.8 %) | 12 (3 %) | 0.977 | 33 (2.9 %) |
| Moderate to severe NDI at 2 years corr. N (%) | 147 (24.4 %) | 85 (22.1 %) | 0.450 | 232 (23.5 %) |
| Cerebral palsy at 2 years corr. N (%) | 36 (6.3 %) | 18 (4.8 %) | 0.394 | 54 (5.7 %) |

#### Main Analyses

**Stratified by Gestational Age**

**Table S3:** Differences in KINDL total score between very preterm born children and their fullterm siblings from FLiP or participants from Ciao Corona, stratified by gestational age (24-27 weeks vs 28-31 weeks). Mean differences denote either differences in health-related quality of life between FLIP very preterm born siblings and their control siblings, or between FLIP very preterm born siblings and controls from Ciao Corona. Negative mean differences indicate that very preterm born children had lower HRQOL than controls. Mean differences and 95% confidence intervals account for either family unit or matching and therefore may not strictly correspond to the means for each group given in the first and second columns.

| Gestational Age | cohort | Preterm | Control | difference | confidence interval |
|---|---|---|---|---|---|
| **Total** | | | | | |
| 24 - 27 weeks | FLiP | 77.6 (10.0) | 80.8 (8.7) | -2.27 | (-4.36 to -0.17) |
| | Ciao Corona | | 77.2 (10.2) | 0.45 | (-1.42 to 2.32) |
| 28 - 31 weeks | FLiP | 78.9 (10.5) | 80.8 (8.7) | -2.29 | (-3.86 to -0.73) |
| | Ciao Corona | | 77.2 (10.2) | 1.72 | ( 0.40 to 3.04) |
| **Physical** | | | | | |
| 24 - 27 weeks | FLiP | 77.4 (17.7) | 82.9 (14.7) | -5.17 | (-9.02 to -1.31) |
| | Ciao Corona | | 77.6 (14.9) | -0.19 | (-3.02 to 2.63) |
| 28 - 31 weeks | FLiP | 82.3 (15.0) | 82.9 (14.7) | -0.44 | (-3.08 to 2.20) |
| | Ciao Corona | | 77.6 (14.9) | 4.74 | ( 2.81 to 6.67) |
| **Emotional** | | | | | |
| 24 - 27 weeks | FLiP | 79.5 (13.0) | 83.6 (11.6) | -3.35 | (-6.10 to -0.61) |
| | Ciao Corona | | 80.5 (14.1) | -0.99 | (-3.55 to 1.56) |
| 28 - 31 weeks | FLiP | 80.8 (14.0) | 83.6 (11.6) | -3.12 | (-5.49 to -0.75) |
| | Ciao Corona | | 80.5 (14.1) | 0.32 | (-1.50 to 2.14) |
| **Self-esteem** | | | | | |
| 24 - 27 weeks | FLiP | 73.9 (13.3) | 76.8 (12.9) | -2.25 | (-5.25 to 0.75) |
| | Ciao Corona | | 72.3 (13.5) | 1.64 | (-0.83 to 4.11) |
| 28 - 31 weeks | FLiP | 73.2 (13.8) | 76.8 (12.9) | -3.68 | (-5.98 to -1.37) |
| | Ciao Corona | | 72.3 (13.5) | 0.87 | (-0.87 to 2.60) |
| **Family** | | | | | |
| 24 - 27 weeks | FLiP | 80.5 (12.6) | 80.3 (12.8) | 1.18 | (-1.60 to 3.96) |
| | Ciao Corona | | 79.1 (12.8) | 1.44 | (-0.91 to 3.79) |
| 28 - 31 weeks | FLiP | 80.2 (13.3) | 80.3 (12.8) | -0.84 | (-2.83 to 1.15) |
| | Ciao Corona | | 79.1 (12.8) | 1.25 | (-0.41 to 2.92) |
| **Friends** | | | | | |
| 24 - 27 weeks | FLiP | 75.5 (14.2) | 78.4 (13.1) | -2.20 | (-5.34 to 0.95) |
| | Ciao Corona | | 77.8 (13.5) | -2.22 | (-4.73 to 0.29) |
| 28 - 31 weeks | FLiP | 76.7 (15.4) | 78.4 (13.1) | -2.07 | (-4.76 to 0.61) |
| | Ciao Corona | | 77.8 (13.5) | -1.06 | (-2.88 to 0.75) |
| **School** | | | | | |
| 24 - 27 weeks | FLiP | 77.5 (16.5) | 82.5 (13.6) | -4.02 | (-7.63 to -0.41) |
| | Ciao Corona | | 75.6 (15.9) | 1.58 | (-1.44 to 4.60) |
| 28 - 31 weeks | FLiP | 80.5 (14.8) | 82.5 (13.6) | -2.11 | (-4.59 to 0.36) |
| | Ciao Corona | | 75.6 (15.9) | 4.81 | ( 2.83 to 6.80) |

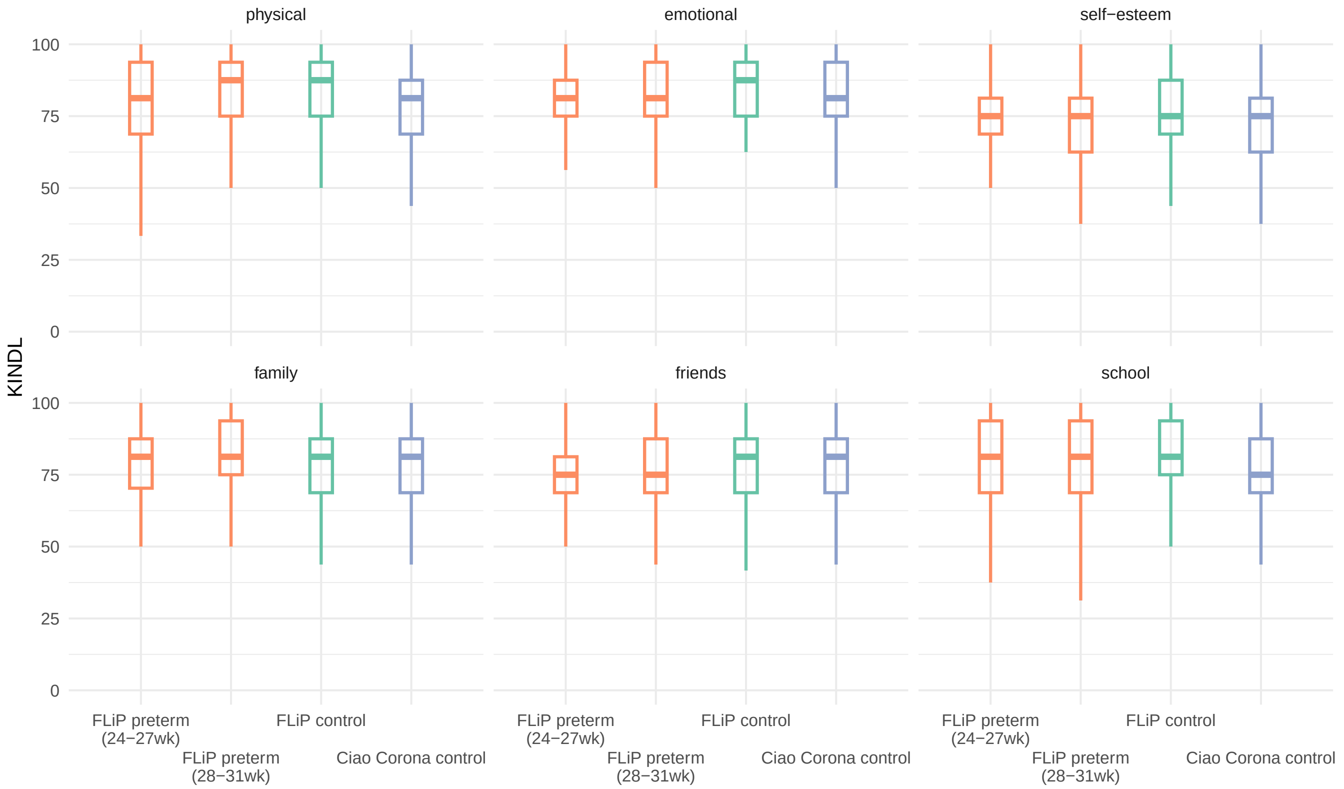

**Figure S2:** KINDL subscales, for very preterm born children (FLiP preterm, stratified by gestational age 24-27 weeks vs 28-31 weeks) and their fullterm born siblings (FLiP control), as well as age, sex and nationality-matched participants from Ciao Corona (Ciao Corona control).

**Stratified by Birthweight**

**Table S4:** Differences in KINDL total score between very preterm born children and their fullterm siblings from FLiP and participants from Ciao Corona, stratified by birthweight ($<$ 1000g vs $\geq$ 1000g). Mean differences denote either differences in health-related quality of life between FLIP very preterm born siblings and their control siblings, or between FLIP very preterm born siblings and controls from Ciao Corona. Mean differences and 95% confidence intervals account for either family unit or matching and therefore may not strictly correspond to the means for each group given in the first and second columns.

| Birthweight | cohort | Preterm | Control | difference | confidence interval |
|---|---|---|---|---|---|
| **Total** | | | | | |
| < 1000g | FLiP | 76.6 (10.4) | 80.8 (8.7) | -3.44 | (-5.48 to -1.41) |
| | Ciao Corona | | 77.2 (10.2) | -0.54 | (-2.27 to 1.19) |
| 1000g + | FLiP | 79.5 (10.1) | 80.8 (8.7) | -1.46 | (-3.05 to 0.12) |
| | Ciao Corona | | 77.2 (10.2) | 2.40 | ( 1.04 to 3.76) |
| **Physical** | | | | | |
| < 1000g | FLiP | 77.0 (16.9) | 82.9 (14.7) | -5.33 | (-8.84 to -1.83) |
| | Ciao Corona | | 77.6 (14.9) | -0.58 | (-3.17 to 2.01) |
| 1000g + | FLiP | 83.0 (15.1) | 82.9 (14.7) | 0.43 | (-2.35 to 3.21) |
| | Ciao Corona | | 77.6 (14.9) | 5.43 | ( 3.43 to 7.43) |
| **Emotional** | | | | | |
| < 1000g | FLiP | 79.8 (13.8) | 83.6 (11.6) | -2.99 | (-5.72 to -0.27) |
| | Ciao Corona | | 80.5 (14.1) | -0.72 | (-3.10 to 1.65) |
| 1000g + | FLiP | 80.8 (13.6) | 83.6 (11.6) | -3.10 | (-5.46 to -0.74) |
| | Ciao Corona | | 80.5 (14.1) | 0.30 | (-1.58 to 2.17) |
| **Self-esteem** | | | | | |
| < 1000g | FLiP | 72.3 (13.4) | 76.8 (12.9) | -3.90 | (-6.73 to -1.06) |
| | Ciao Corona | | 72.3 (13.5) | -0.01 | (-2.278 to 2.25) |
| 1000g + | FLiP | 74.0 (13.8) | 76.8 (12.9) | -2.71 | (-5.08 to -0.33) |
| | Ciao Corona | | 72.3 (13.5) | 1.72 | (-0.076 to 3.52) |
| **Family** | | | | | |
| < 1000g | FLiP | 80.0 (12.6) | 80.3 (12.8) | -0.01 | (-2.65 to 2.63) |
| | Ciao Corona | | 79.1 (12.8) | 0.94 | (-1.21 to 3.09) |
| 1000g + | FLiP | 80.5 (13.3) | 80.3 (12.8) | -0.19 | (-2.20 to 1.82) |
| | Ciao Corona | | 79.1 (12.8) | 1.52 | (-0.22 to 3.25) |
| **Friends** | | | | | |
| < 1000g | FLiP | 73.6 (16.0) | 78.4 (13.1) | -4.51 | (-7.79 to -1.22) |
| | Ciao Corona | | 77.8 (13.5) | -4.17 | (-6.53 to -1.82) |
| 1000g + | FLiP | 77.9 (14.2) | 78.4 (13.1) | -0.93 | (-3.46 to 1.60) |
| | Ciao Corona | | 77.8 (13.5) | 0.15 | (-1.68 to 1.98) |
| **School** | | | | | |
| < 1000g | FLiP | 76.4 (16.8) | 82.5 (13.6) | -5.20 | (-8.73 to -1.67) |
| | Ciao Corona | | 75.6 (15.9) | 0.67 | (-2.08 to 3.41) |
| 1000g + | FLiP | 81.3 (14.3) | 82.5 (13.6) | -1.61 | (-4.02 to 0.81) |
| | Ciao Corona | | 75.6 (15.9) | 5.63 | ( 3.58 to 7.68) |

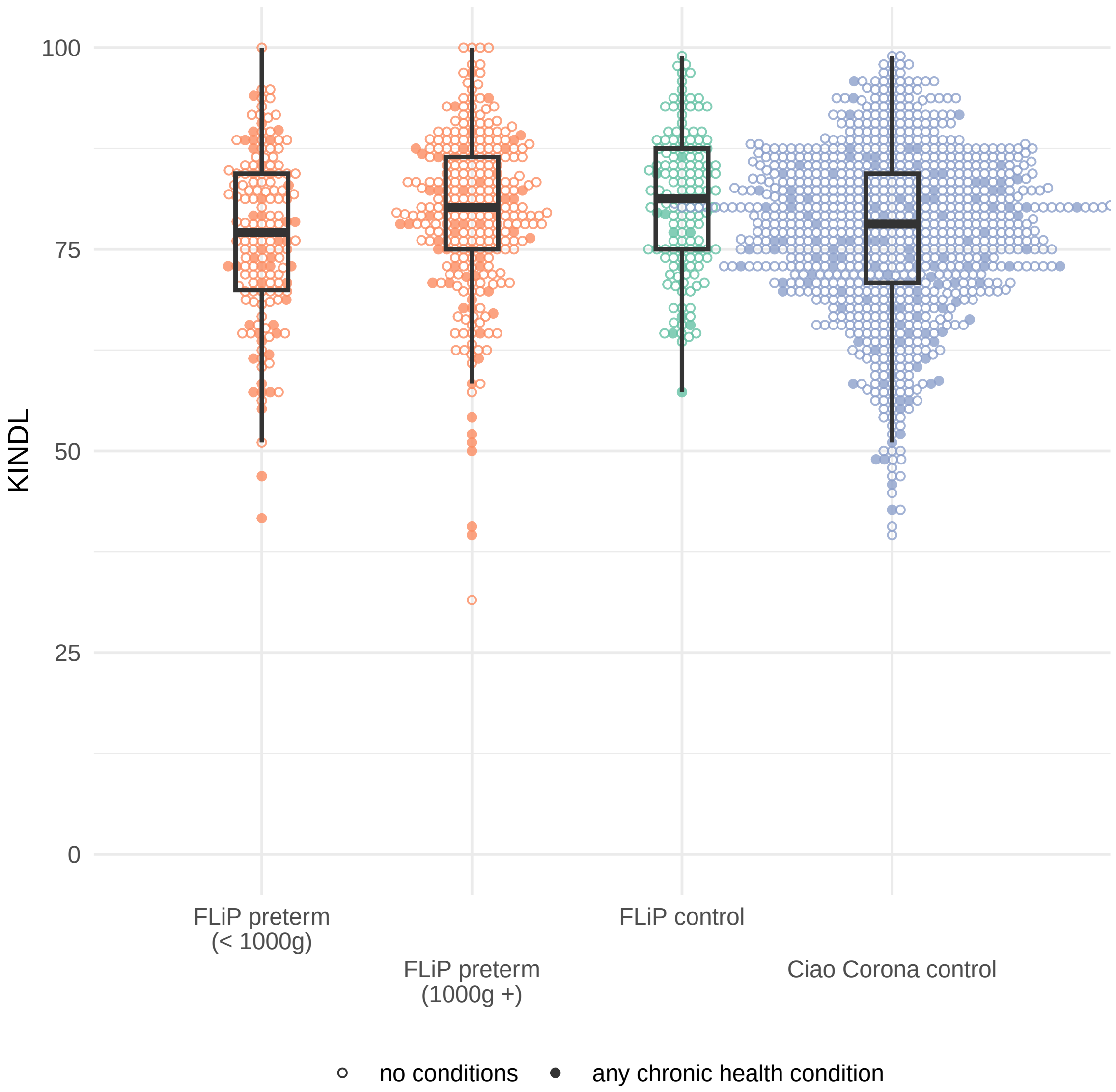

**Figure S3:** KINDL total score, for very preterm born children (FLiP preterm, stratified by birthweight $<$ 1000g vs $\geq$ 1000g) and their fullterm born siblings (FLiP control), as well as age, sex and nationality-matched participants from Ciao Corona (Ciao Corona control). Solid circles indicate participants without chronic health conditions, while empty diamonds indicate those with any chronic health condition.

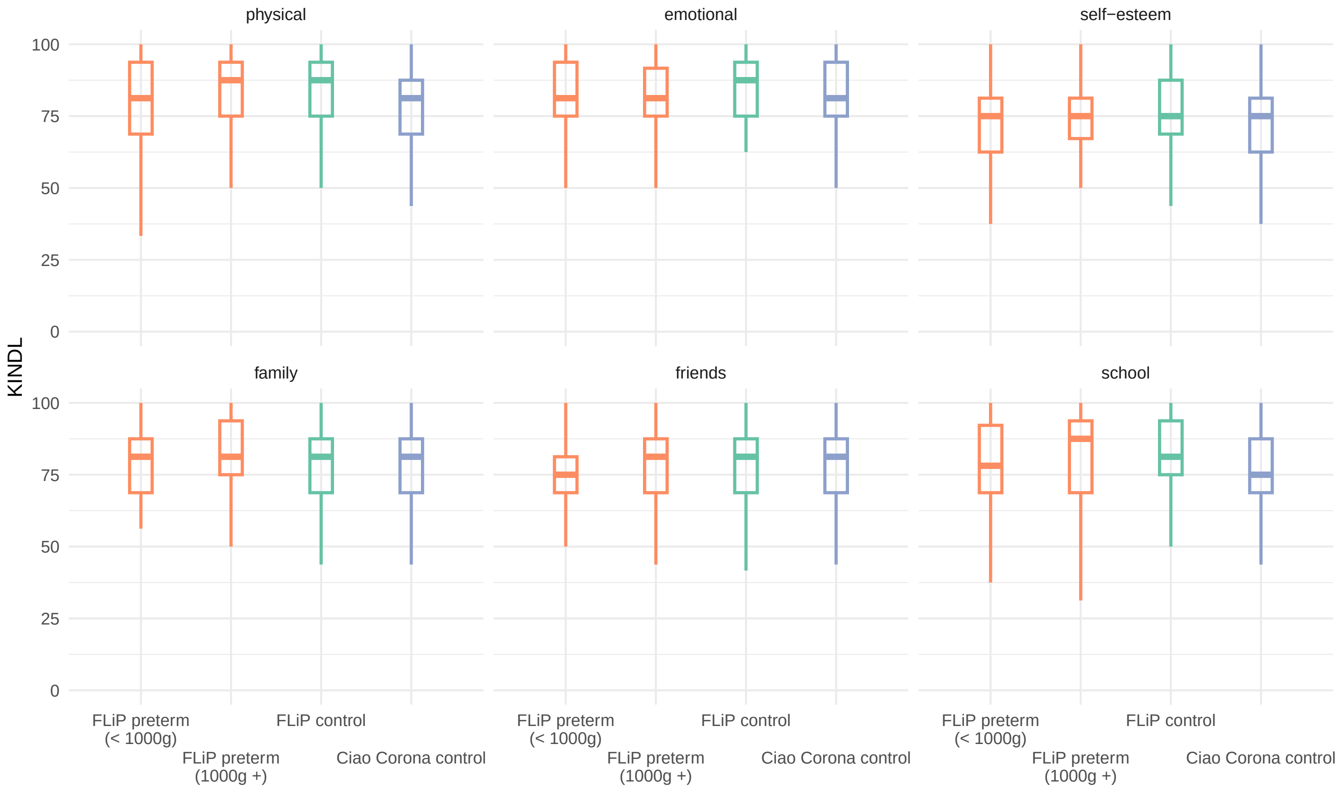

**Figure S4:** KINDL subscales, for very preterm born children (FLiP preterm, stratified by birthweight: <1000g vs >1000g) and their fullterm born siblings (FLiP control), as well as age, sex and nationality-matched participants from Ciao Corona (Ciao Corona control).

**All very preterm born children, regardless of gestational age or birthweight**

**Table S5:** Differences in KINDL total score between very preterm born children and their fullterm siblings from FLiP and participants from Ciao Corona. Mean differences denote either differences in health-related quality of life between FLIP very preterm born siblings and their control siblings, or between FLIP very preterm born siblings and controls from Ciao Corona. Mean differences and 95% confidence intervals account for either family unit or matching and therefore may not strictly correspond to the means for each group given in the first and second columns.

| cohort | Preterm | Control | difference | confidence interval |
|---|---|---|---|---|
| **Total** | | | | |
| FLiP | 78.5 (10.3) | 80.8 (8.7) | -2.09 | (-3.56 to -0.62) |
| Ciao Corona | | 77.2 (10.2) | 1.35 | (0.19 to 2.51) |
| **Physical** | | | | |
| FLiP | 80.9 (16.0) | 82.9 (14.7) | -1.34 | (-4.02 to 1.34) |
| Ciao Corona | | 77.6 (14.9) | 3.28 | (1.53 to 5.03) |
| **Emotional** | | | | |
| FLiP | 80.4 (13.7) | 83.6 (11.6) | -3.16 | (-5.31 to -1.00) |
| Ciao Corona | | 80.5 (14.1) | -0.08 | (-1.67 to 1.52) |
| **Self-esteem** | | | | |
| FLiP | 73.4 (13.6) | 76.8 (12.9) | -3.22 | (-5.39 to -1.06) |
| Ciao Corona | | 72.3 (13.5) | 1.10 | (-0.42 to 2.61) |
| **Family** | | | | |
| FLiP | 80.3 (13.1) | 80.3 (12.8) | -0.20 | (-2.04 to 1.64) |
| Ciao Corona | | 79.1 (12.8) | 1.31 | (-0.16 to 2.77) |
| **Friends** | | | | |
| FLiP | 76.4 (15.0) | 78.4 (13.1) | -2.21 | (-4.68 to 0.26) |
| Ciao Corona | | 77.8 (13.5) | -1.40 | (-3.01 to 0.20) |
| **School** | | | | |
| FLiP | 79.7 (15.3) | 82.5 (13.6) | -2.49 | (-4.87 to -0.11) |
| Ciao Corona | | 75.6 (15.9) | 3.94 | (2.19 to 5.69) |

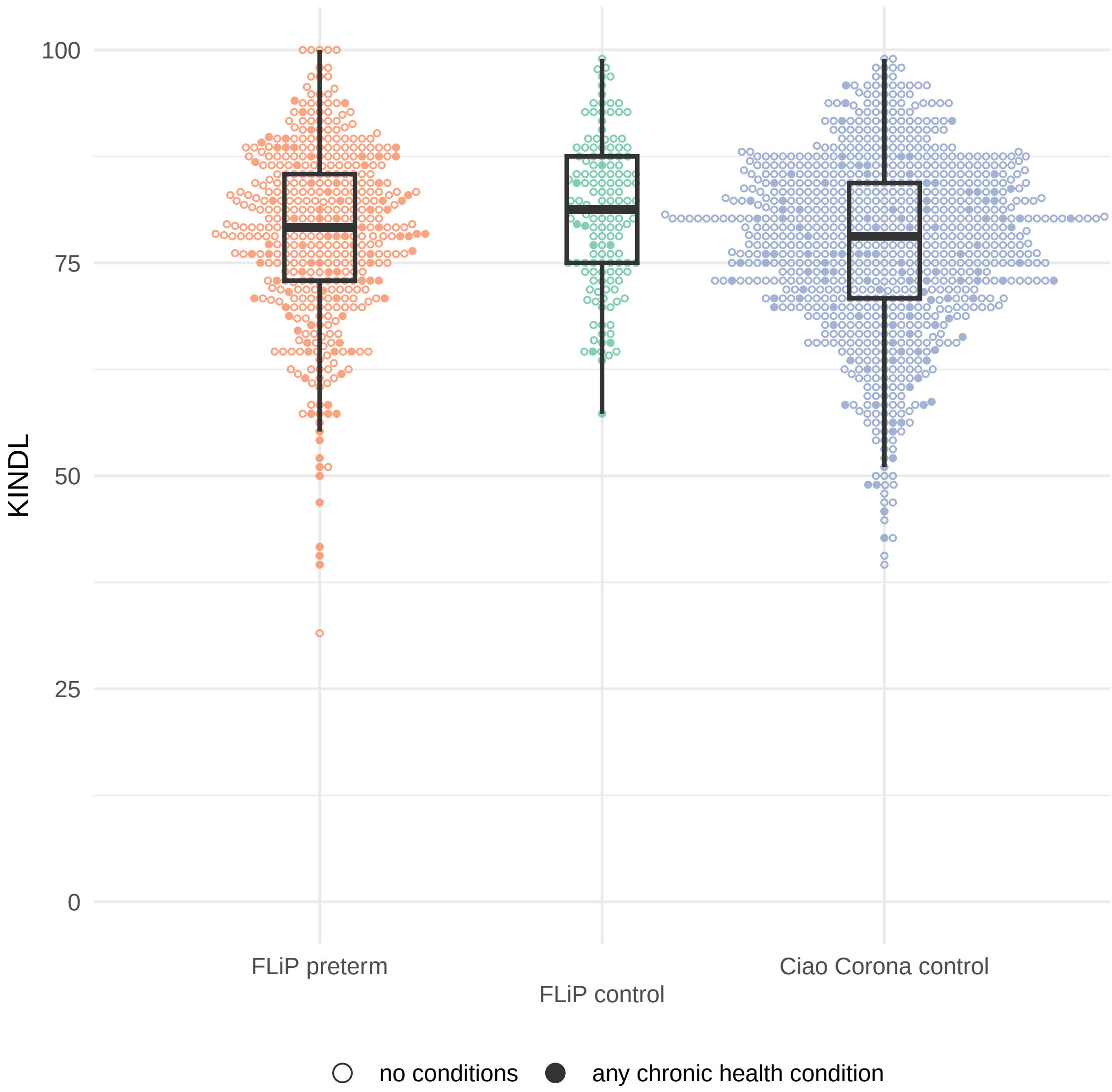

**Figure S5:** KINDL total score, for very preterm born children (FLiP preterm) and their fullterm born siblings (FLiP control), as well as age, sex and nationality-matched participants from Ciao Corona (Ciao Corona control). Solid circles indicate participants without chronic health conditions, while empty diamonds indicate those with any chronic health condition.

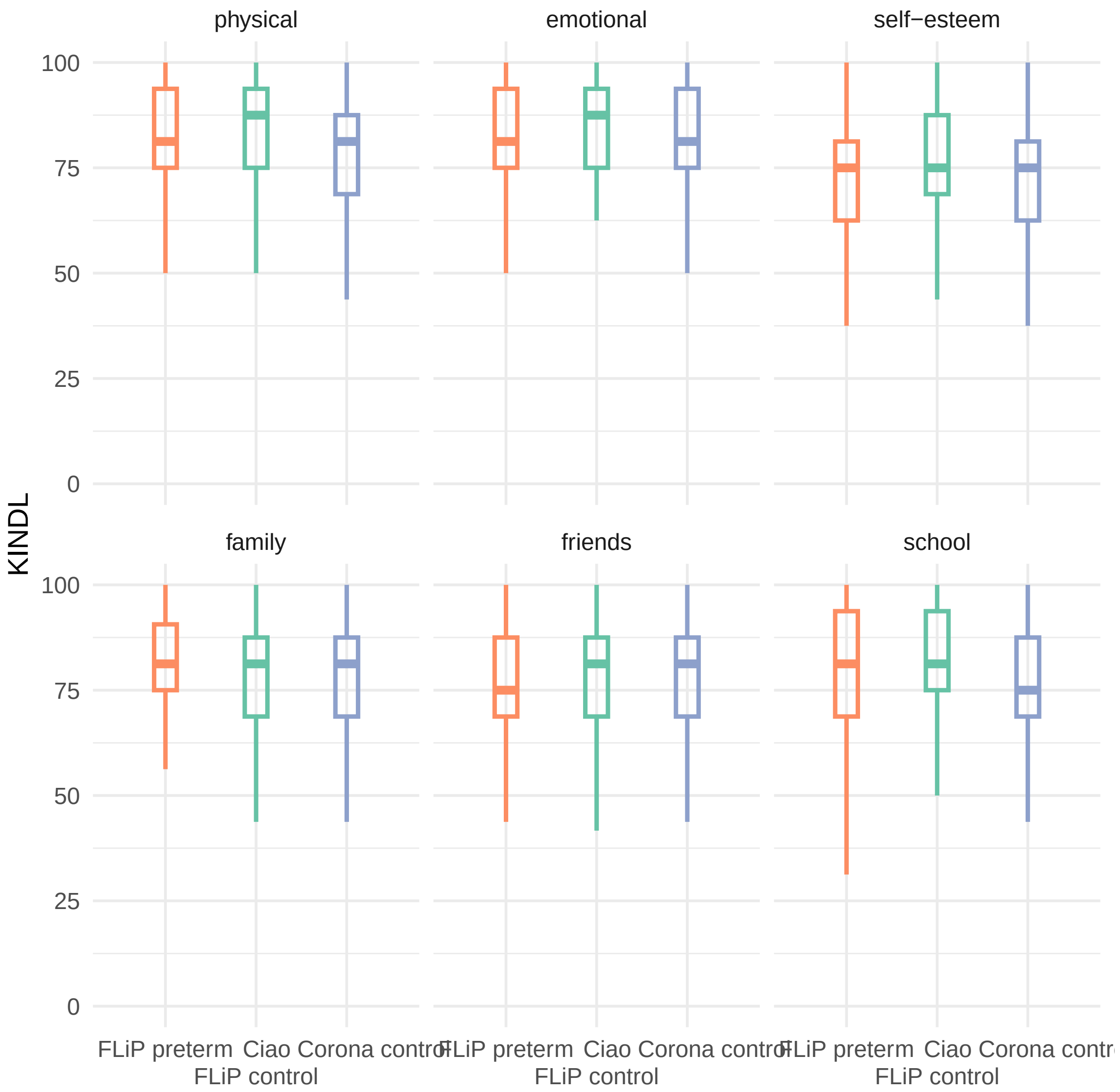

**Figure S6:** KINDL subscales, for very preterm born children (FLiP preterm) and their fullterm born siblings (FLiP control), as well as age, sex and nationality-matched participants from Ciao Corona (Ciao Corona control).

**Table of potential determinants**

**Table S6:** Table of potential determinants of health-related quality of life in FLiP among very preterm born children and adolescents, by age group.

| **Characteristic** | **Overall**, N = 442 | **5-9**, N = 227 | **10+**, N = 215 | **p-value** |
|---|---|---|---|---|
| KINDL total score | 79 (73, 85) | 80 (74, 85) | 79 (72, 86) | 0.11 |
| sex (male) | 236 (53%) | 116 (51%) | 120 (56%) | 0.3 |
| overweight | 56 (13%) | 26 (12%) | 30 (15%) | 0.5 |
| Unknown | 27 | 17 | 10 | |
| non-Swiss nationality | 66 (15%) | 39 (17%) | 27 (13%) | 0.2 |
| Unknown | 1 | 1 | 0 | |
| socio-economic status | 5 (3, 6) | 5 (3, 6) | 5 (3, 6) | 0.14 |
| Unknown | 12 | 1 | 11 | |
| unemployed | 103 (24%) | 50 (22%) | 53 (25%) | 0.4 |
| Unknown | 6 | 1 | 5 | |
| siblings | | | | 0.6 |
| 0 | 91 (21%) | 49 (22%) | 42 (20%) | |
| 1 | 229 (53%) | 120 (54%) | 109 (52%) | |
| 2+ | 114 (26%) | 54 (24%) | 60 (28%) | |
| Unknown | 8 | 4 | 4 | |
| smoking | | | | 0.007 |
| no | 349 (79%) | 174 (77%) | 175 (81%) | |
| outside | 84 (19%) | 52 (23%) | 32 (15%) | |
| in the house | 9 (2.0%) | 1 (0.4%) | 8 (3.7%) | |
| pets | 169 (38%) | 59 (26%) | 110 (51%) | <0.001 |
| physical activity (hrs / day) | 0.7 (0.5, 1.0) | 0.7 (0.5, 1.0) | 0.7 (0.5, 1.0) | 0.4 |
| Unknown | 5 | 2 | 3 | |
| screen time (hrs / day) | 1.1 (0.6, 2.0) | 0.8 (0.5, 1.1) | 1.9 (1.0, 2.6) | <0.001 |
| Unknown | 11 | 6 | 5 | |
| Participation in sports outside of school | 319 (73%) | 167 (74%) | 152 (72%) | 0.7 |
| Unknown | 3 | 0 | 3 | |
| Participation in music lessons or activities | 128 (29%) | 58 (26%) | 70 (33%) | 0.085 |
| Unknown | 3 | 0 | 3 | |
| Participation in scouts or similar | 48 (11%) | 20 (8.8%) | 28 (13%) | 0.14 |
| Unknown | 3 | 0 | 3 | |
| Gestational age | | | | 0.006 |
| 24-27 wks | 130 (29%) | 80 (35%) | 50 (23%) | |
| 28-31 wks | 312 (71%) | 147 (65%) | 165 (77%) | |
| birthweight | | | | 0.2 |
| <1000g | 158 (36%) | 87 (38%) | 71 (33%) | |
| 1000+g | 284 (64%) | 140 (62%) | 144 (67%) | |
| BPD | 55 (12%) | 29 (13%) | 26 (12%) | 0.8 |
| chronic non-respiratory conditions | 67 (15%) | 32 (14%) | 35 (16%) | 0.5 |
| chronic respiratory conditions | 24 (5.4%) | 9 (4.0%) | 15 (7.0%) | 0.2 |
| cerebral palsy | 33 (7.5%) | 12 (5.3%) | 21 (9.8%) | 0.073 |
| therapy | | | | 0.032 |
| no therapy | 307 (69%) | 145 (64%) | 162 (75%) | |
| 1-2 | 116 (26%) | 70 (31%) | 46 (21%) | |
| 3+ | 19 (4.3%) | 12 (5.3%) | 7 (3.3%) | |
| assistive devices | 15 (3.4%) | 7 (3.1%) | 8 (3.7%) | 0.7 |
| respiratory symptoms affecting daily life | 99 (22%) | 54 (24%) | 45 (21%) | 0.5 |

[1] Median (IQR); n (%)

[2] [illegible]

#### Sensitivity Analyses

Sensitivity analyses included

a) excluding participants with chronic health conditions;

  - Because very preterm born children were much more likely to report chronic non-respiratory conditions, respiratory conditions or cerebral palsy than fullterm children (24% vs 8% in our sample), we also examined a subset of FLiP participants that did not report any chronic health condition. In this subset, preterm children had on average a 1.3 point lower total KINDL score that their fullterm siblings (95% CI -2.8 to 0.2) and 2.0 points higher KINDL total score (0.7 to 3.2) than controls in Ciao Corona (**Table S7, Figures S7 and S8**).

b) restricting to preterm born children with control siblings;

  - To ensure comparable groups with respect to family characteristics, including socioeconomic status and general level of physical activity, we restricted to those families with both preterm and fullterm siblings as a sensitivity analysis, leaving 119 preterm children and 119 controls (**Table S8**). Among those families, very preterm born children had on average 2.0 points lower KINDL total score than their fullterm born siblings (95% CI -3.7 to -0.3, **Table S9, Figure S9**).

c) stratification by age; and

  - We noted similar patterns in both younger and older children (**Figures S10 and S11, Table S10**). The mean difference in KINDL total score between preterm and controls was -2.1 (95% CI -4.0 to -0.1) among children 5-9 years of age, and -1.7 (-4.2 to 0.7) among those 10 and older.

d) adjusting for SES.

  - SES was directly influenced by family unit, and likely contributes to HRQOL. We therefore considered models with and without SES. The two models showed similar fit (BIC 4194 vs 4201), and the estimates were of very similar magnitude (-2.1 [-3.6 to -0.6] in both cases).

e) accounting for family unit using fixed rather than random effects.

  - The random effects and fixed effects models gave very similar results, both for all participants in FLiP (**Figure S12 and Table S11**) and for subgroups stratified by gestational age (**Figure S13 and Table S12**).

None of the sensitivity analyses led to a different conclusion regarding HRQOL in very preterm born children compared to their peers.

**a) excluding participants with chronic health conditions**

**Table S7:** Differences in KINDL total score between very preterm born children, their fullterm born siblings, and Ciao Corona participants (stratified by presence of any chronic health conditions). In FLiP, 104 of very preterm born children had chronic health conditions (24%), while n = 12 of their control siblings did (8%). Among controls from Ciao Corona, 122 (14%) reported chronic health conditions. Mean differences denote either differences in health-related quality of life between FLIP very preterm born siblings and their control siblings, or between FLIP very preterm born siblings and controls from Ciao Corona. Mean differences and 95% confidence intervals account for either family unit or matching and therefore may not strictly correspond to the means for each group given in the first and second columns.

| chronic health conditions | cohort | Preterm | Control | difference | confidence interval |
|---|---|---|---|---|---|
| **Total** | | | | | |
| any chronic health condition | FLiP | 74.5 (12.2) | 73.8 (8.7) | -4.50 | (-10.39 to 1.39) |
| | Ciao Corona | | 73.8 (10.3) | 0.69 | (-2.26 to 3.65) |
| no chronic health conditions | FLiP | 79.7 (9.4) | 81.4 (8.5) | -1.30 | (-2.82 to 0.21) |
| | Ciao Corona | | 77.7 (10.1) | 1.99 | (0.74 to 3.23) |
| **Physical** | | | | | |
| any chronic health condition | FLiP | 77.0 (17.2) | 75.5 (10.8) | 0.58 | (-9.44 to 10.60) |
| | Ciao Corona | | 74.0 (14.3) | 3.17 | (-0.96 to 7.30) |
| no chronic health conditions | FLiP | 82.1 (15.4) | 83.5 (14.9) | -0.75 | (-3.59 to 2.09) |
| | Ciao Corona | | 78.2 (15.0) | 3.87 | (1.93 to 5.81) |
| **Emotional** | | | | | |
| any chronic health condition | FLiP | 78.4 (15.2) | 77.6 (17.2) | -2.36 | (-10.72 to 5.99) |
| | Ciao Corona | | 77.3 (14.7) | 1.31 | (-2.63 to 5.24) |
| no chronic health conditions | FLiP | 81.0 (13.1) | 84.1 (10.9) | -3.14 | (-5.38 to -0.89) |
| | Ciao Corona | | 81.1 (13.9) | -0.09 | (-1.81 to 1.64) |
| **Self-esteem** | | | | | |
| any chronic health condition | FLiP | 68.1 (16.7) | 70.3 (10.0) | -5.38 | (-14.27 to 3.52) |
| | Ciao Corona | | 68.7 (13.3) | -0.52 | (-4.48 to 3.43) |
| no chronic health conditions | FLiP | 75.0 (12.2) | 77.3 (13.0) | -1.93 | (-4.03 to 0.16) |
| | Ciao Corona | | 72.9 (13.5) | 2.03 | (0.38 to 3.67) |
| **Family** | | | | | |
| any chronic health condition | FLiP | 78.9 (14.1) | 76.0 (12.5) | -1.92 | (-9.49 to 5.66) |
| | Ciao Corona | | 75.9 (13.8) | 3.25 | (-0.43 to 6.93) |
| no chronic health conditions | FLiP | 80.7 (12.7) | 80.6 (12.8) | 0.37 | (-1.50 to 2.24) |
| | Ciao Corona | | 79.6 (12.5) | 1.15 | (-0.44 to 2.75) |
| **Friends** | | | | | |
| any chronic health condition | FLiP | 71.4 (15.6) | 69.6 (14.9) | 1.05 | (-8.22 to 10.33) |
| | Ciao Corona | | 73.2 (15.7) | -1.80 | (-5.85 to 2.24) |
| no chronic health conditions | FLiP | 77.9 (14.5) | 79.2 (12.7) | -1.28 | (-3.84 to 1.28) |
| | Ciao Corona | | 78.5 (13.0) | -0.57 | (-2.30 to 1.16) |
| **School** | | | | | |
| any chronic health condition | FLiP | 72.6 (18.0) | 72.2 (19.2) | -3.29 | (-12.48 to 5.89) |
| | Ciao Corona | | 73.8 (16.8) | -1.44 | (-6.08 to 3.20) |
| no chronic health conditions | FLiP | 81.9 (13.7) | 83.5 (12.7) | -1.52 | (-4.05 to 1.00) |
| | Ciao Corona | | 75.9 (15.8) | 5.77 | (3.89 to 7.66) |

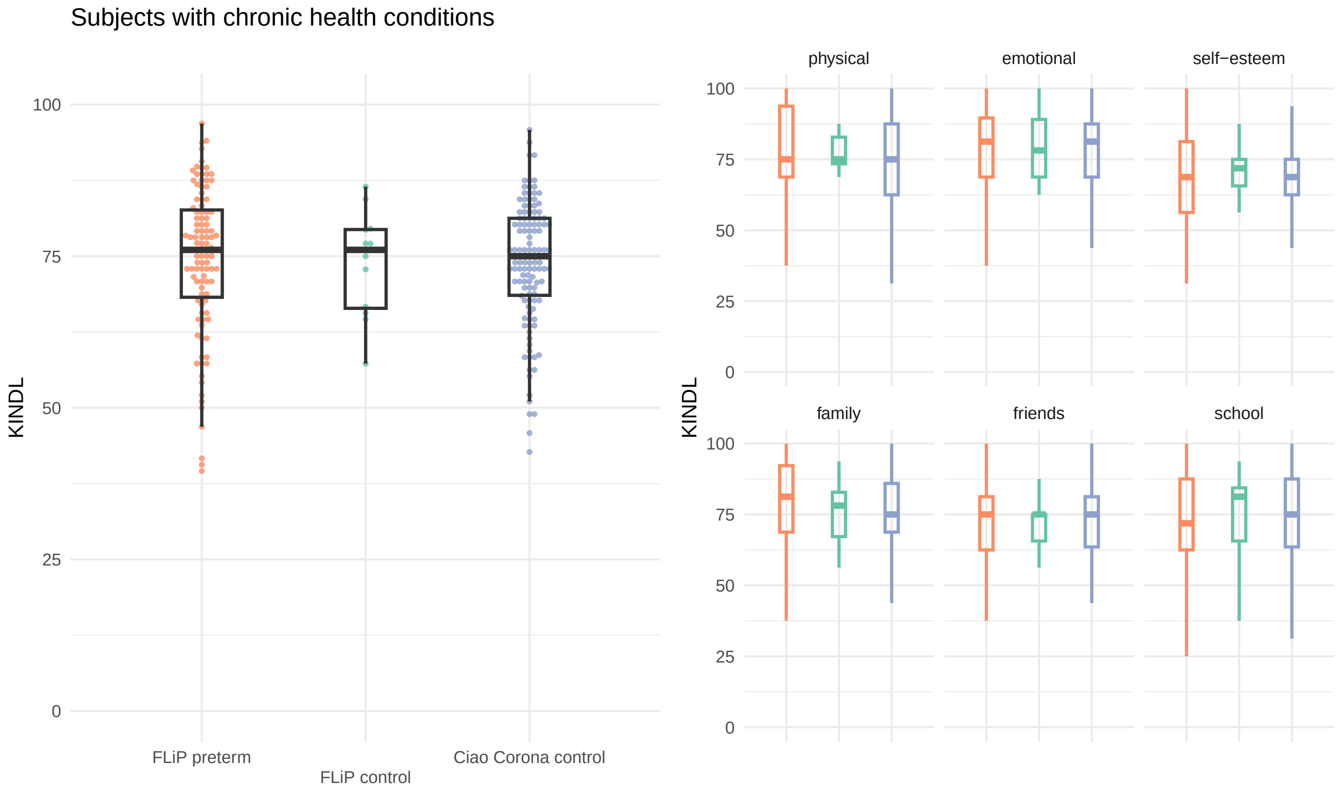

**Figure S7:** KINDL total score and its subscales, for preterm children and their fullterm siblings (with any chronic health condition)

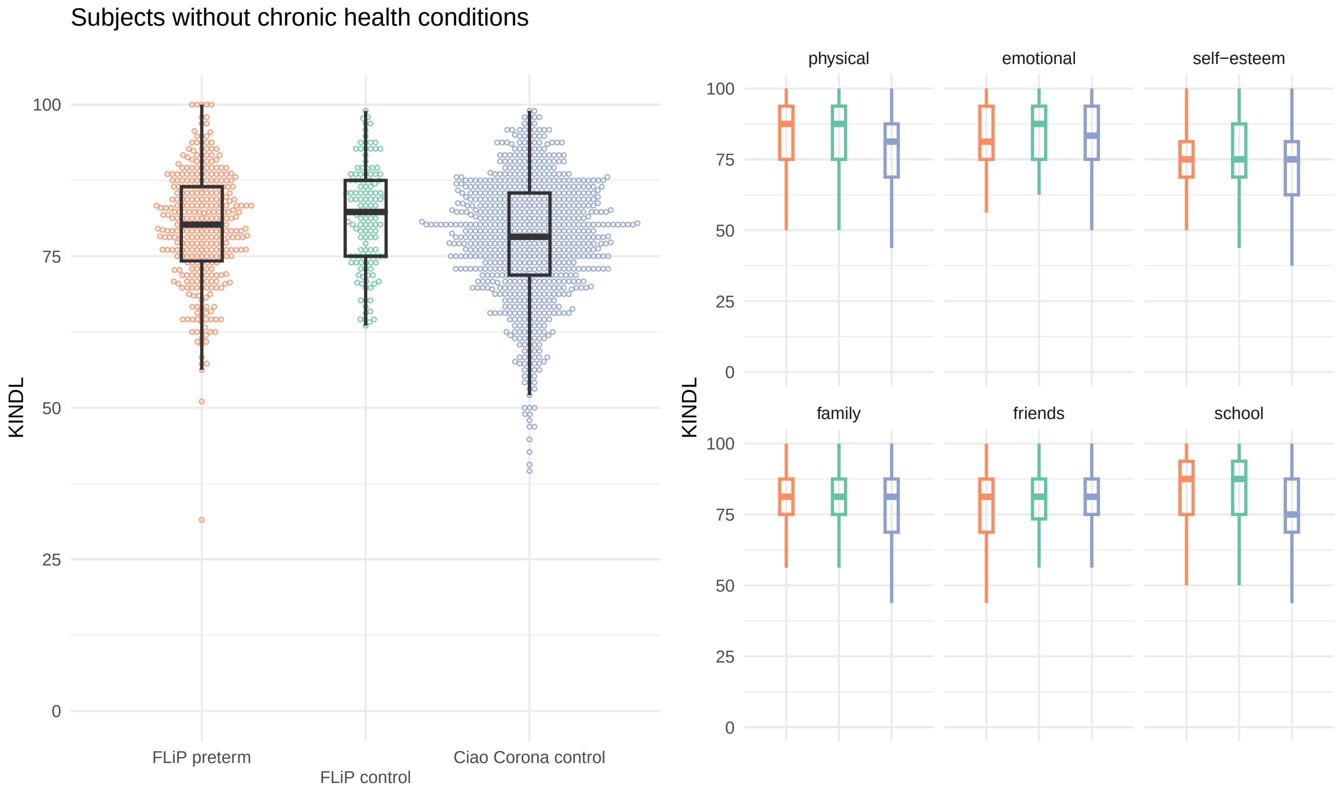

**Figure S8:** KINDL total score and its subscales, for preterm children and their fullterm siblings (with no chronic health conditions)

##### b) restricting to preterm born children with control siblings

**Table S8:** Differences in KINDL total score between preterm children and their fullterm siblings (only matched siblings).

| KINDL | Preterm | Control | difference | confidence interval |
|---|---|---|---|---|
| total | 78.5 (10.5) | 80.6 (8.9) | -1.99 | (-3.66 to -0.32) |
| physical | 82.1 (15.0) | 81.9 (15.6) | 0.19 | (-3.08 to 3.45) |
| emotional | 80.5 (13.7) | 83.6 (11.5) | -3.02 | (-5.66 to -0.38) |
| self-esteem | 72.6 (14.0) | 76.3 (13.1) | -3.26 | (-5.79 to -0.74) |
| family | 80.1 (12.8) | 80.9 (12.5) | -0.52 | (-2.52 to 1.48) |
| friends | 75.8 (15.3) | 78.2 (13.2) | -2.46 | (-5.36 to 0.45) |
| school | 79.4 (14.7) | 82.5 (13.7) | -2.67 | (-5.58 to 0.23) |

**Table S9:** Key characteristics of preterm and control participants in full data (preterm children with included control siblings)

| **Characteristic** | **FLiP preterm**, N = 119 | **FLiP control**, N = 119 |
|---|---|---|
| age (years) | 11 (5 - 16) | 10 (5 - 19) |
| sex (male) | 62 (52%) | 71 (61%) |
| gestational age (weeks) | | |
| 24-27 wks | 25 (21%) | |
| 28-31 wks | 94 (79%) | |
| birthweight | | |
| <1000g | 28 (24%) | |
| 1000+g | 91 (76%) | |
| multiple gestation | 28 (24%) | |
| socio-economic status | 5 [3, 6] | 5 [3, 6] |
| non-Swiss nationality | 15 (13%) | 12 (10%) |
| moderate to severe BPD | 15 (13%) | |
| coughing / wheezing restrict daily activities | 4 (3.4%) | 1 (0.8%) |
| any chronic health condition | 31 (26%) | 10 (8.4%) |
| chronic non-respiratory conditions | 21 (18%) | 9 (7.6%) |
| chronic respiratory conditions | 9 (7.6%) | 2 (1.7%) |
| cerebral palsy | 8 (6.7%) | 1 (0.8%) |
| physical activity (hours per week) | 0.71 [0.57, 1.00] | 0.75 [0.57, 1.14] |

[1] Median (Range); n (%); Median [IQR]

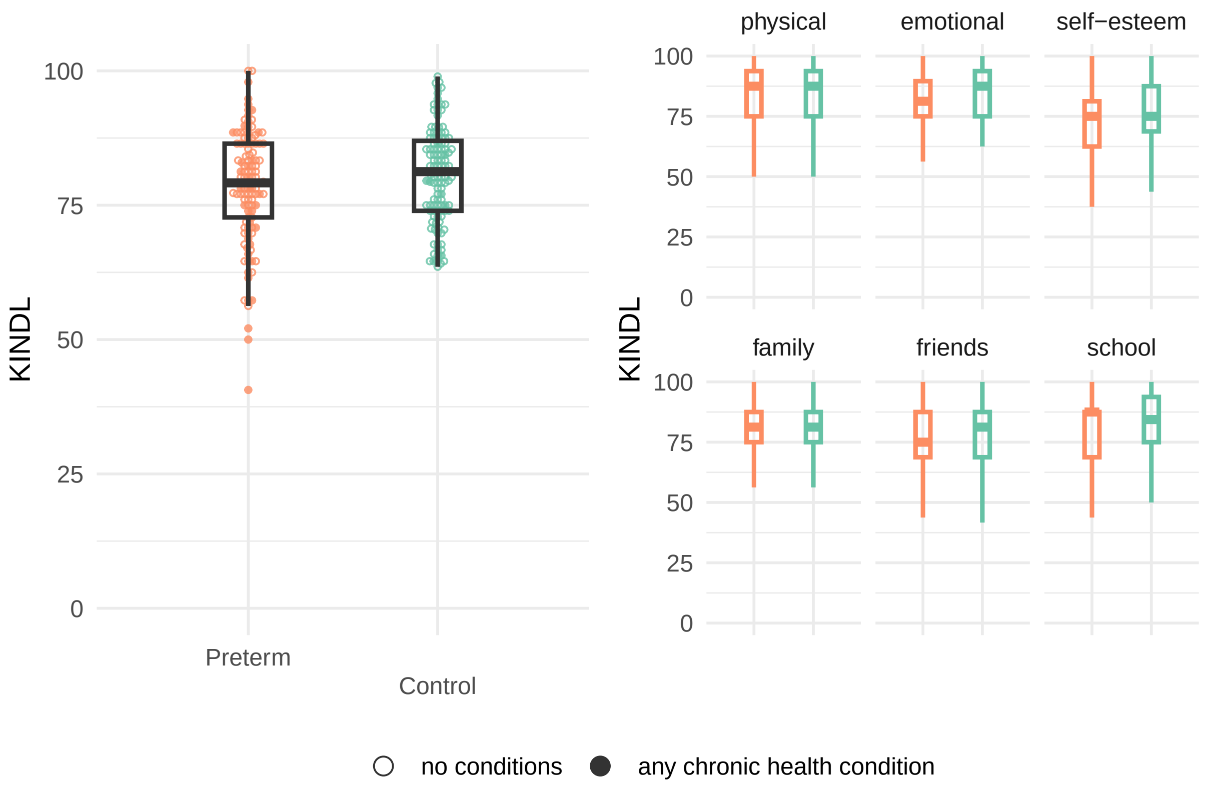

**Figure S9:** KINDL total score and its subscales, for preterm children and their fullterm siblings (matched siblings only)

**c) stratification by age**

**Table S10:** Differences in KINDL total score between very preterm born children and their fullterm siblings from FLiP and participants from Ciao Corona, by age group (5-9, 10+). Mean differences, 95% confidence intervals and p-values account for either family unit or matching and therefore may not strictly correspond to the means for each group given in the first and second columns.

| age | cohort | Preterm | Control | difference | confidence interval |
|---|---|---|---|---|---|
| **Total** | | | | | |
| 5-9 | FLiP | 79.6 (8.7) | 82.1 (8.0) | -2.06 | (-3.97 to -0.14) |
| | Ciao Corona | | 79.8 (9.0) | -0.07 | (-1.49 to 1.35) |
| 10+ | FLiP | 77.3 (11.7) | 78.8 (9.5) | -1.73 | (-4.18 to 0.73) |
| | Ciao Corona | | 75.3 (10.7) | 2.00 | (0.24 to 3.75) |
| **Physical** | | | | | |
| 5-9 | FLiP | 81.2 (16.1) | 85.5 (13.1) | -4.14 | (-7.74 to -0.54) |
| | Ciao Corona | | 80.9 (13.6) | 0.37 | (-2.05 to 2.79) |
| 10+ | FLiP | 80.5 (16.0) | 79.1 (16.1) | 2.09 | (-2.01 to 6.20) |
| | Ciao Corona | | 75.2 (15.4) | 5.29 | (2.83 to 7.76) |
| **Emotional** | | | | | |
| 5-9 | FLiP | 81.0 (12.6) | 84.3 (11.0) | -3.31 | (-6.14 to -0.49) |
| | Ciao Corona | | 82.3 (13.0) | -1.19 | (-3.29 to 0.92) |
| 10+ | FLiP | 79.8 (14.8) | 82.6 (12.5) | -3.23 | (-6.65 to 0.19) |
| | Ciao Corona | | 79.3 (14.7) | 0.52 | (-1.83 to 2.87) |
| **Self-esteem** | | | | | |
| 5-9 | FLiP | 74.9 (12.1) | 78.8 (11.4) | -3.51 | (-6.35 to -0.68) |
| | Ciao Corona | | 74.4 (13.0) | 0.51 | (-1.48 to 2.51) |
| 10+ | FLiP | 71.8 (15.0) | 73.8 (14.4) | -2.76 | (-6.45 to 0.94) |
| | Ciao Corona | | 70.8 (13.7) | 1.05 | (-1.17 to 3.27) |
| **Family** | | | | | |
| 5-9 | FLiP | 79.0 (12.2) | 79.7 (12.8) | -0.43 | (-3.13 to 2.27) |
| | Ciao Corona | | 79.0 (12.2) | 0.08 | (-1.91 to 2.07) |
| 10+ | FLiP | 81.7 (13.8) | 81.1 (12.7) | -0.22 | (-2.97 to 2.54) |
| | Ciao Corona | | 79.1 (13.2) | 2.59 | (0.48 to 4.71) |
| **Friends** | | | | | |
| 5-9 | FLiP | 77.9 (12.5) | 79.3 (12.0) | -1.28 | (-4.22 to 1.66) |
| | Ciao Corona | | 79.4 (12.5) | -1.60 | (-3.67 to 0.47) |
| 10+ | FLiP | 74.8 (17.1) | 77.0 (14.5) | -2.29 | (-6.43 to 1.84) |
| | Ciao Corona | | 76.6 (14.1) | -1.74 | (-4.15 to 0.66) |
| **School** | | | | | |
| 5-9 | FLiP | 84.4 (12.4) | 85.2 (11.9) | 0.03 | (-2.93 to 2.99) |
| | Ciao Corona | | 82.5 (13.1) | 1.85 | (-0.30 to 4.00) |
| 10+ | FLiP | 74.5 (16.5) | 78.7 (14.9) | -3.50 | (-7.29 to 0.29) |
| | Ciao Corona | | 70.7 (16.0) | 3.85 | (1.20 to 6.50) |

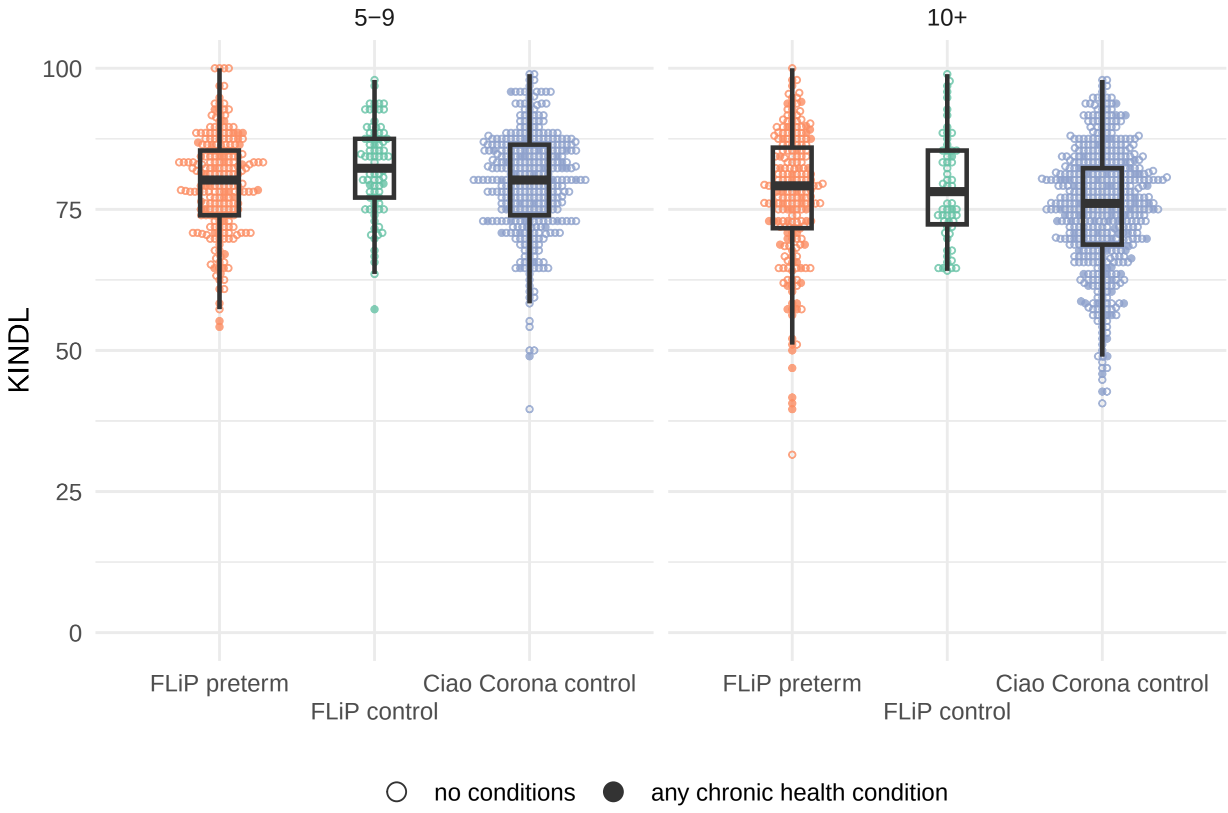

**Figure S10:** KINDL total score in very preterm born children and fullterm born controls from FLiP, as well as controls from Ciao Corona, by age group. Solid circles indicate participants without chronic health conditions, while empty circles indicate those with any chronic health condition.

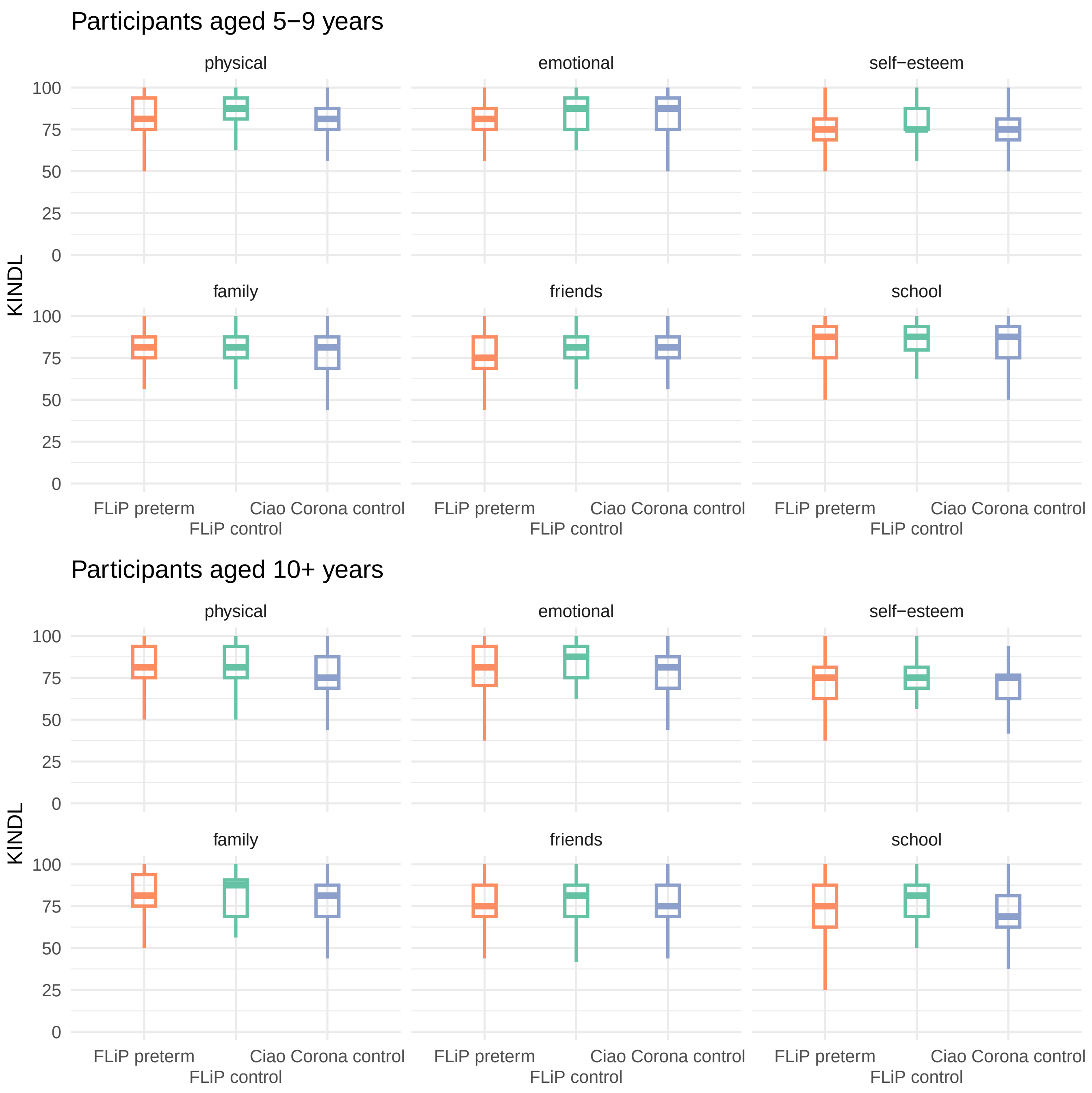

**Figure S11:** KINDL subscales in very preterm born children and fullterm born controls from FLiP, as well as controls from Ciao Corona, by age group.

##### d) adjusting for SES

SES was directly influenced by family unit, and likely contributes to HRQOL. We therefore considered models with and without SES. The two models showed similar fit (BIC 4194 vs 4201), and the estimates were of very similar magnitude (-2.1 [-3.6 to -0.6] in both cases).

```
sm2 <- sm %>%
  ungroup() %>%
  select(kindl_total, group, family_id, SES_Score_total) %>%
  filter(complete.cases(.))
mod_noSES <- lmer(kindl_total ~ group + (1 | family_id), data = sm2)
tidy(mod_noSES, conf.int = TRUE) %>%
  filter(term == "groupPreterm") %>%
  select(term, estimate, conf.low, conf.high, p.value) %>%
  mutate(BIC = BIC(mod_noSES),
         AIC = AIC(mod_noSES))
```

```
## # A tibble: 1 x 7
##   term         estimate conf.low conf.high p.value   BIC   AIC
##   <chr>           <dbl>    <dbl>     <dbl>   <dbl> <dbl> <dbl>
## 1 groupPreterm    -2.10    -3.59    -0.613 0.00581 4194. 4176.
```

```
mod_SES <- lmer(kindl_total ~ group + SES_Score_total + (1 | family_id), data = sm2)
tidy(mod_SES, conf.int = TRUE) %>%
  filter(term == "groupPreterm") %>%
  select(term, estimate, conf.low, conf.high, p.value) %>%
  mutate(BIC = BIC(mod_SES),
         AIC = AIC(mod_SES))
```

```
## # A tibble: 1 x 7
##   term         estimate conf.low conf.high p.value   BIC   AIC
##   <chr>           <dbl>    <dbl>     <dbl>   <dbl> <dbl> <dbl>
## 1 groupPreterm    -2.09    -3.58    -0.605 0.00600 4201. 4179.
```

**e) accounting for family unit using fixed rather than random effects**

The random effects and fixed effects models gave very similar results, both for all participants in FLiP (**Figure S12 and Table S11**) and for subgroups stratified by gestational age (**Figure S13 and Table S12**).

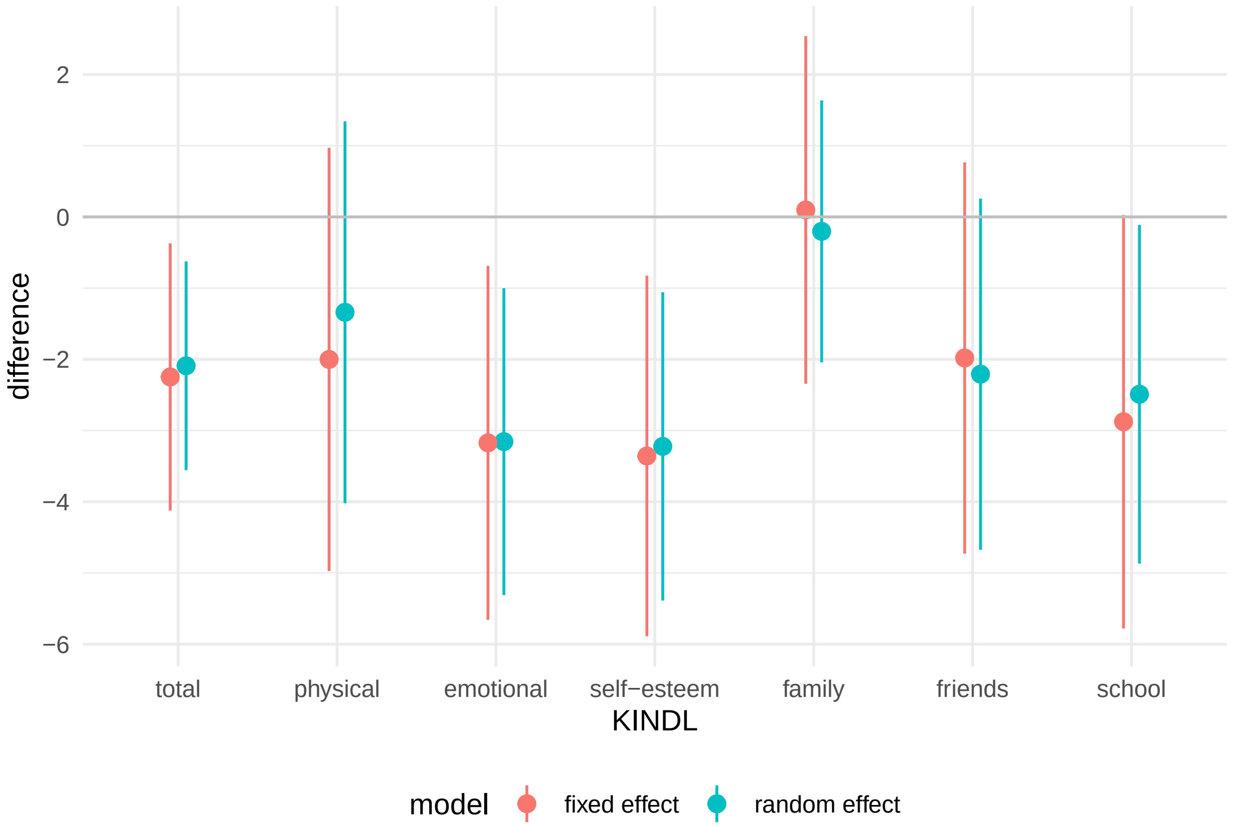

**Figure S12:** Results from models comparing FLiP participants, accounting for family unit using fixed effects.

**Table S11:** Differences in KINDL total score between very preterm born children and their fullterm siblings from FLiP. Results from models accounting for family unit as 1) random effect, or 2) fixed effect are shown. Mean differences and 95% confidence intervals are shown.

| KINDL | random effect | fixed effect |
|---|---|---|
| total | -2.09 (-3.56 to -0.62) | -2.25 (-4.12 to -0.37) |
| physical | -1.34 (-4.02 to 1.34) | -2.00 (-4.97 to 0.97) |
| emotional | -3.16 (-5.31 to -1.00) | -3.17 (-5.66 to -0.69) |
| self-esteem | -3.22 (-5.39 to -1.06) | -3.36 (-5.89 to -0.82) |
| family | -0.203 (-2.04 to 1.64) | 0.098 (-2.34 to 2.54) |
| friends | -2.21 (-4.68 to 0.26) | -1.98 (-4.73 to 0.77) |
| school | -2.49 (-4.87 to -0.11) | -2.88 (-5.78 to 0.027) |

**Table S12:** Differences in KINDL total score between very preterm born children and their fullterm siblings from FLiP, by gestational age. Results from models accounting for family unit as 1) random effect, or 2) fixed effect are shown. Mean differences and 95% confidence intervals are shown.

| KINDL | Gestational Age | random effect | fixed effect |
|---|---|---|---|
| total | 24 - 27 weeks | -2.27 (-4.36 to -0.17) | 0.94 (-2.99 to 4.88) |
| total | 28 - 31 weeks | -2.29 (-3.86 to -0.73) | -2.65 (-4.48 to -0.82) |
| physical | 24 - 27 weeks | -5.169 (-9.02 to -1.31) | 1.119 (-6.99 to 9.23) |
| physical | 28 - 31 weeks | -0.441 (-3.08 to 2.20) | -0.046 (-3.35 to 3.26) |
| emotional | 24 - 27 weeks | -3.35 (-6.10 to -0.61) | -0.79 (-5.84 to 4.26) |
| emotional | 28 - 31 weeks | -3.12 (-5.49 to -0.75) | -3.42 (-6.40 to -0.43) |
| self-esteem | 24 - 27 weeks | -2.25 (-5.25 to 0.75) | -0.18 (-6.07 to 5.72) |
| self-esteem | 28 - 31 weeks | -3.68 (-5.98 to -1.37) | -3.89 (-6.74 to -1.03) |
| family | 24 - 27 weeks | 1.18 (-1.60 to 3.96) | 3.79 (-0.96 to 8.53) |
| family | 28 - 31 weeks | -0.84 (-2.83 to 1.15) | -1.57 (-3.84 to 0.71) |
| friends | 24 - 27 weeks | -2.20 (-5.34 to 0.95) | 0.56 (-5.53 to 6.65) |
| friends | 28 - 31 weeks | -2.07 (-4.76 to 0.61) | -3.24 (-6.69 to 0.20) |
| school | 24 - 27 weeks | -4.02 (-7.63 to -0.41) | -2.08 (-9.44 to 5.27) |
| school | 28 - 31 weeks | -2.11 (-4.59 to 0.36) | -2.18 (-5.21 to 0.85) |

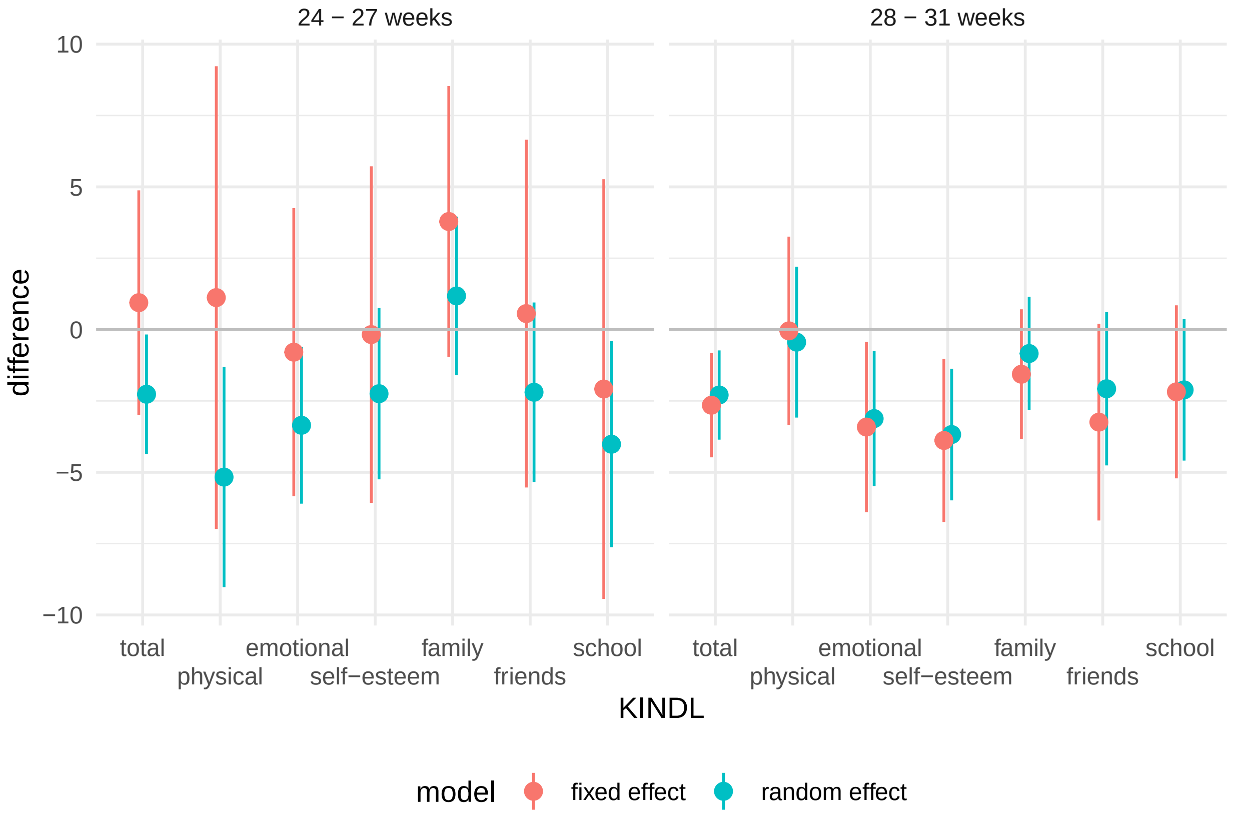

**Figure S13:** Results from models comparing FLiP participants, by age group, accounting for family unit using fixed effects.

#### Computational Details

- R version: R version 4.4.1 (2024-06-14)
- Base packages: grid, stats, graphics, grDevices, utils, datasets, methods, base
- Other packages: consort 1.2.1, kableExtra 1.4.0, ggparty 1.0.0, partykit 1.2.20, mvtnorm 1.2.4, libcoin 1.0.10, patchwork 1.2.0, ggdag 0.2.12, gtsummary 1.7.2, lmerTest 3.1.3, lme4 1.1.35.5, Matrix 1.7.0, broom.mixed 0.2.9.5, broom 1.0.5, ggbeeswarm 0.7.2, lubridate 1.9.3, forcats 1.0.0, stringr 1.5.1, dplyr 1.1.4, purrr 1.0.2, readr 2.1.5, tidyr 1.3.1, tibble 3.2.1, ggplot2 3.5.1, tidyverse 2.0.0

This document was generated on 2024-09-04 at 10:37.
